# Good practice in the provision of care for people living with dementia in nursing homes: a systematic review

**DOI:** 10.1101/2024.03.04.24302868

**Authors:** Laura Behan, Michael P O’Brien, Paul Dunbar, Niall McGrane, Aileen Keane, Carol Grogan, Laura M Keyes

**Author notes:** Corresponding Author: Laura Keyes.

## Abstract

**Background:** The number of people living with dementia across the world is rising, and there is a high and ever increasing proportion of people with dementia living in nursing homes. It is increasingly important that care provision in these services accounts for the specific needs of this cohort. Manifestations of dementia are modifiable with high quality dementia specific care, as such, we need to understand what good practice looks like specifically in these settings.

**Aim:** To synthesise empirical research to identify the characteristics of quality in the provision of care for people living with dementia in nursing homes.

**Methods:** Four electronic academic databases were searched: Business Complete, CINAHL, MEDLINE and APA PsychInfo. Qualitative, quantitative or mixed-methods studies published between the years of 2020-2023, that aimed to identify determinants of high quality care for people with dementia living in nursing homes, were included. Themes relating to good practices were identified and narratively summarised. Vignettes illustrating good practice were constructed from the perspective of a provider, a staff member, a resident and a family member.

**Results:** After screening of 3,356 records, 30 articles were included. This included 16 qualitative descriptive studies, 11 cross-sectional studies, 2 mixed methods studies, and 1 cohort study. Sixteen themes were identified: determinants of care quality, outcomes, person-centred care, cultural impact, care planning, meaningful engagement, eating and meals, the role of family members, restrictive practices, psychotropic medications, activities, materials, health care, end of life care, staffing and staff training.

**Conclusions:** The literature identifies what constitutes high quality care for residents with dementia in nursing homes. These findings will guide those delivering care in nursing homes in their daily work and in the implementation of quality improvement processes. It will also direct regulators, policymakers and researchers when conducting future work in this important area.

## Introduction

Dementia is a syndrome, chronic and progressive in nature. It is the broad term used for various age-related neurodegenerative disorders that impact a person’s cognitive skills such as language, memory, judgement, and learning capacity.(1) The number of people living with dementia across the world is rising with numbers projected to increase from current figures of 50 million people to, 152 million people in 2050.(2) At least 70% of nursing home residents in the United Kingdom and over 50% of nursing home residents in the United States of America live with dementia.(3, 4)

In 2012, the World Health Organisation (WHO) and the Alzheimer’s Disease International emphasised the public health priority status of dementia.(1) One of the key messages from the report is for countries to include dementia on their public health agendas; this requires sustained action and coordination at international, national, regional and local levels.(1) In 2013, the G8 Dementia Summit was convened to generate an international response to dementia, and agreed on a list of commitments with a primary aim to identify a cure or disease-modifying therapy for dementia by 2025. They developed a coordinated international action plan for dementia research, which would be delivered by the newly formed World Dementia Council.(5) Although this event was an important marker in dementia care, the majority of the commitments were not specific, measurable, or time bound, and did not have any reporting or accountability mechanisms in place.(6)

In 2017, the Lancet Commission on Dementia Prevention, Intervention, and Care met to consolidate up-to-date knowledge on the prevention and management of dementia, particularly with the health systems in high-income countries.(7) The Commission was reconvened and updated in 2020(8) and presented the latest evidence on intervention and care for dementia. These reports highlighted that while the underlying illness of dementia is not curable, many of its manifestations are now known to be manageable, and can be modifiable with good dementia care. Good dementia care considers the person as a whole and encompasses medical, social, and supportive care; it should be personalised to unique individual and cultural needs, preferences, and priorities, and should incorporate support for family carers.(7, 8) Furthermore, the Alzheimer’s Disease International’s 2022 World Alzheimer Report on diagnosis and post-diagnosis support recognises the importance of person-centred care; the need for support for carers; the evolving role of clinicians through the post-diagnostic journey in care planning, treatment, and monitoring; the importance of a robust post-diagnostic care model for enabling a cohesive relationship between people living with dementia, carers, and health-care professionals; and the need for healthcare professionals to receive more training in dementia care.(9) These reports, while providing overarching guidelines on interventions and care, do not however focus on specific individual components of high care quality within nursing home settings.

Given the high and ever increasing proportion of people with dementia in nursing homes, and the current emphasis for countries to have a sustained action plan for the provision of care, we need to understand what good practice looks like specifically in these settings. This systematic review aimed to synthesise the characteristics of quality in the provision of dementia care in nursing homes.

## Methods

A systematic review was conducted of studies that used qualitative, quantitative or mixed-methods approaches to identify determinants of high quality care for people with dementia living in nursing homes. We take “dementia” to mean any form of cognitive disorders, dementia, Alzheimer’s, vascular or frontotemporal disorder. “Nursing home” was defined, for the purposes of this review, as any residential institution that provides older people, who are permanent residents, round the clock care from the staff in the nursing home, which may or may not have included qualified nursing staff. Good practices were anything associated with quality of care and could also be aspects of the organisation or the environment. Barriers or facilitators to successful implementation of quality care were also considered.

### Eligibility Criteria

The PICo (Population, Interest, Context) framework was used to determine study eligibility. Literature meeting the following criteria was sought for inclusion:

- Population: older people living with dementia
- Interest: good practices in the provision of quality care
- Context: nursing homes

Facilities providing acute healthcare and or inpatient rehabilitation services only, and residential services specifically for people of all ages with intellectual and or physical disabilities were excluded.

Any analytic study design – experimental, quasi-experimental or observational – was deemed eligible for inclusion, as were mixed-methods studies that included an analytical component. Reviews, abstracts, opinion pieces, discussion papers, letters to the editor, magazine articles, blog posts, and information materials were excluded.

Only articles published between the years of 2020-2023 were included. The rationale for this time frame included the publication of the Lancet Commission on Dementia Prevention, Intervention, and Care in 2017, updated in 2020. This consolidated up-to-date knowledge on the prevention and management of dementia.(8) This time limit was also selected to ensure that the most up-to-date evidence was prioritised. In addition, only reports available in English were included.

### Search Strategy

Relevant search words were identified following a scoping search of literature using strings combined with the “AND” operator. Search terms were used to identify, retrieve and evaluate literature from academic databases. Four electronic academic databases were searched in April 2023: Business Complete, CINAHL, MEDLINE and APA PsychInfo. Identification of known key articles within the search results confirmed the sensitivity of the search. Reference list checks of included articles was conducted to identify any further articles for inclusion. The search terms are provided in the supplementary section.

### Study selection

All articles were imported into Covidence(10) and duplicates were deleted. Titles, abstracts and full texts were then screened. Titles and abstracts were screened by one reviewer (LB). Full-text review was carried out independently by two reviewers (LB and LMK) with conflicts resolved through discussion and consensus.

### Data extraction

Data extraction was performed by LB. A standardised data extraction table was constructed using the following headings: author; jurisdiction; study design; aim; characteristics of sample; overview of results, and criteria or elements of what constitutes high quality of care.

### Quality assessment

Critical appraisal of the studies was performed to ensure the trustworthiness and relevance of data. The quality of quasi-experimental studies, qualitative studies and cross-sectional studies were critically appraised using standard critical appraisal tools prepared by the Joanna Briggs Institute (JBI).(11) Because JBI does not provide an appraisal tool for mixed methods designs, the Mixed Methods Appraisal Tool by Hong et al., 2018 was used to appraise these studies.(12) Critical appraisal was completed independently by two reviewers (LB and MOB). Discrepancies were resolved through consensus between the authors.

### Data synthesis

A narrative synthesis of findings was compiled and themes relating to good practices were identified and described. Due to the heterogeneous nature of study designs, outcomes and outcome measures, a meta-analysis was not possible. Four vignettes illustrating good practice were constructed by two researchers (LB and MOB) following immersion in the identified themes. The vignettes were written from the perspective of a provider, a staff members, a resident and a family member.

## Results

An initial search of the empirical literature identified 3,356 records. Following removal of 1,291 duplicates, 2,065 records remained for title and abstract screening. During screening, 1,995 irrelevant records were excluded based on our eligibility criteria and eights reports were inaccessible, leaving 62 for full-text review. Upon full-text review, 32 reports were excluded on account of being the wrong setting (n=3), wrong outcomes (n=22), wrong study design (n=5) or wrong population (n=2). (Figure 1 PRISMA flow diagram). No records were identified from reference list checks.

**Figure 1:**
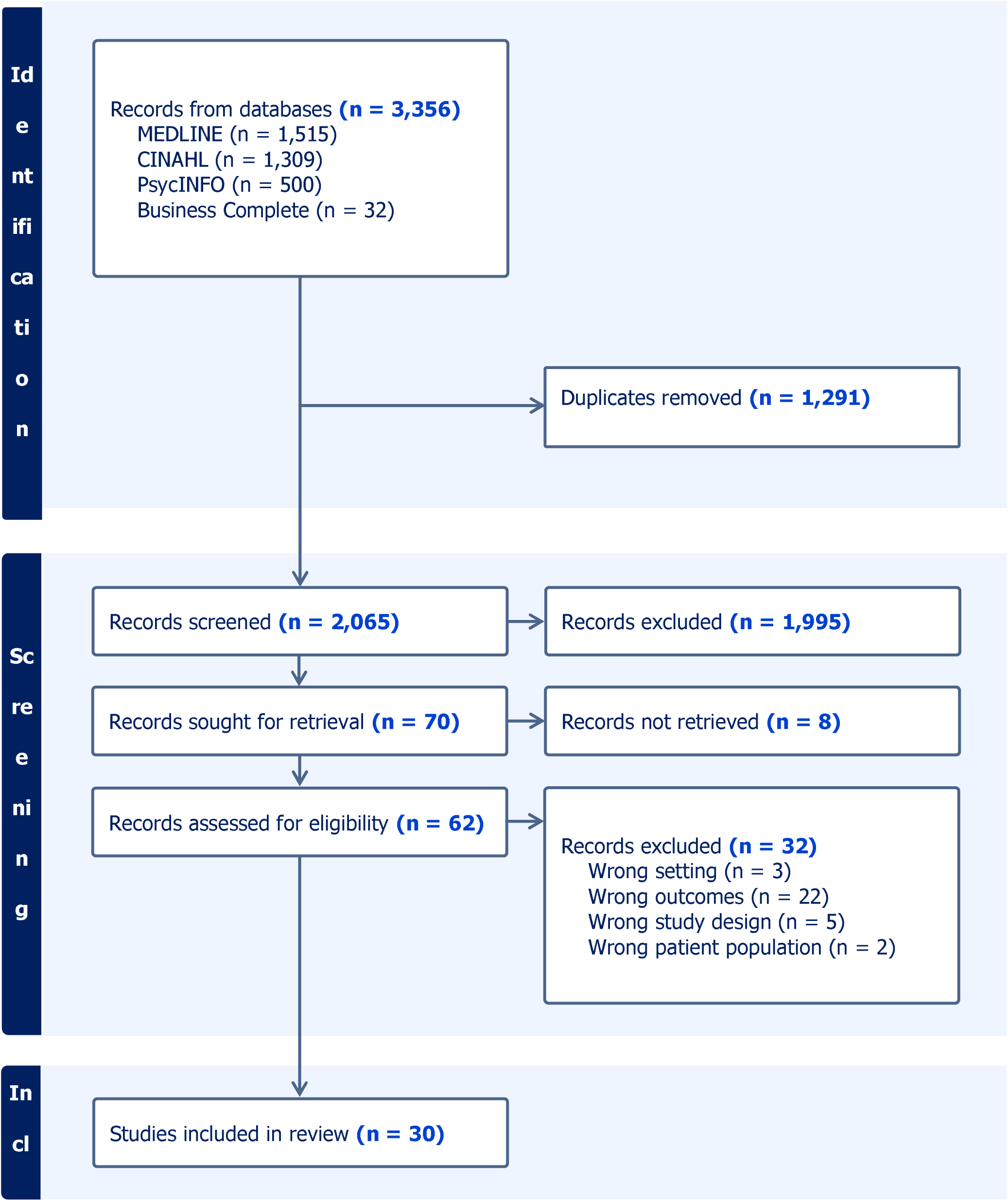
PRISMA flow diagram.

### Study Characteristics

Of the 30 studies included in the review, 16 were qualitative descriptive studies,(13, 14, 15, 16, 17, 18, 19, 20, 21, 22, 23, 24, 25, 26, 27, 28) 11 were cross-sectional studies,(28, 29, 30, 31, 32, 33, 34, 35, 36, 37, 38) 2 were mixed methods studies,(39, 40) and 1 was a cohort study.(41) Participants included people living with dementia, relatives, staff working in nursing homes, experts in dementia care, administrators and lawyers. Settings were described as nursing homes (n=20), residential care facilities (n=3), dementia care unit (n=3), care homes (n=2), assisted living facility (n=1) and hospital or nursing homes specialised in palliative care (n=1). Studies were conducted in Australia (n = 8),(15, 16, 17, 26, 28, 30, 40, 42) USA (n = 6),(14, 20, 24, 33, 37, 43) UK (n = 5),(19, 20, 21, 39, 42) Norway (n = 4),(18, 23, 27, 29) the Netherlands (n = 2),(31, 35) Denmark (n = 2),(22, 25) New Zealand (n=1)(42) Sweden (n = 2),(34, 36) Canada (n = 1),(32) Northern Ireland (n = 1),(31) Indonesia (n=1)(13), South Africa (n=1),(42) and Malaysia (n = 1).(41) Two of these studies were conducted in multiple jurisdictions.(31, 42) The characteristics of included studies are set out in Supplementary table 1.

### Quality appraisal of included studies

For the quantitative studies, there were mixed findings in terms of quality. Most followed best practice in terms of defining their samples, identifying and using appropriate statistical methods. However, a considerable number of studies did not identify confounding factors or employ strategies to control for confounding factors.

Most qualitative and mixed methods studies were of good quality. There was generally a congruence between the methods employed by the researchers and their objectives, data collection methods, analysis and interpretation of results. Common weaknesses in these studies tended to be a lack of congruity between the stated philosophical perspective and the research methodology, and the absence of a statement locating the authors culturally and theoretically in the research.

### Themes

Sixteen themes were identified through the systematic review and are presented in table 1. These themes include: determinants of care quality, outcomes, person-centred care, cultural impact, care planning, meaningful engagement, eating and meals, the role of family members, restrictive practices, psychotropic medications, activities, materials, health care, end of life care, staffing and staff training. The findings from the review promote a person-centred approach to care where residents are involved in all decisions around their care and support and staff work closely with the resident living with dementia, their caregivers and their relatives. Effective communication with the resident is key to meaningful engagement; this can be supported by staff understanding the importance of getting to know residents, learning their communication patterns and effective verbal and non-verbal communication techniques. The findings also promote individualised care planning, training in dementia centred practices, involvement of resident in activities, promoting cultural appropriate care, and the reduction of anti-psychotic medication and use of restraint. Challenges exist in providing high quality care for residents with dementia in nursing homes and include staff shortages, high workloads, insufficient resources and insufficient dementia specific staff training.

**Table 1:** Good practice in providing care for people living with dementia in nursing homes, themes identified through systematic review of the literature (2020 to 2023).

| Theme | Description | Studies addressing theme | Key findings |
| --- | --- | --- | --- |
| Determinants of care quality | This theme presents how quality of care is measured or determined within the included studies. | 7(13, 15, 16, 30, 37, 38, 39) | 1. Measures used to quantify care quality included the percentage of residents who were prescribed antipsychotic medication(30, 37, 38) or benzodiazepines(38); and the percentage who experienced adverse events, were physically restrained, or who lost weight.(30, 37, 39)<br>2. Scaled scores used to measure quality of care included person-centred care scale score and quality of care scale score.(30)<br>3. Quality of care was determined by how staff treated the resident, assistance at meals,(30) deprivation of liberty, use of cognitive enhancing medications, staff friendliness, and if meal-times were pleasurable.(39)<br>4. Quality nursing care linked to the nurse–patient therapeutic relationship and included meeting the physical, spiritual and emotional needs of each person, providing comfort, reaching the person within the disease, and making a difference to each person every day.(13, 16, 39)<br>5. Characteristics of a well-performing nursing home include good accountability, communication, staff-resident-family connections, and trust in the care provided.(15) |
| Outcomes | Monitoring outcomes of quality care is considered good practice in the provision of care. | 3(18, 30, 39) | 1. Outcomes for determining high quality of care were pain, depression, BMI, food and fluid intake, agitated behaviours, frailty, engagement, positive behaviours and QOL scores.(30)<br>2. Resident's autonomy, social relationships, joy of life and meaningful activities were also measured.(18)<br>3. Enhanced personhood was assessed through engagement with one's local community, fulfilment of one's spiritual needs, and provision and engagement in meaningful modes of life and activity.(39) |
| Person-centred care | Person-centred care is a term used to describe a standard of care that places the person at the heart of care delivery. It is considered best practice for care of people with dementia. | 8(14, 17, 18, 28, 32, 35, 41, 44) | 1. The term "person-centred dementia care" were distinguished from "person-centred care" as being more concerned with connections, relationships, and the role care staff play in ensuring connections.(28)<br>2. Person-centred care was shown to have a positive impact on quality of care, enhanced resident well-being, and better staff satisfaction and retention.(35, 41, 44)<br>3. Residents with dementia were found to not always receive person-centred dementia care.(35, 41, 44)<br>4. Staff reported challenges in implementing person-centred care including demand on staff time, staff shortages forcing staff to prioritise task-based duties and difficulties shifting mind sets.(17, 28, 32)<br>5. Person-centred practice should not be an "add on" that new long-term providers "buy into" but should infiltrate every activity within an organisation and all staff should demonstrate it.(14, 41)<br>6. Improved person-centred care was achieved through providing greater autonomy to residents, purposeful relationship building, acknowledging identities(28, 31), focusing on behavioural support, placing person-centeredness above protocols, insuring training on person-centred culture, valuing implementation flexibility,(14) and individual care planning.(28)<br>7. Improvements require working closely with the person living |

| Theme | Description | Studies addressing theme | Key findings |
| --- | --- | --- | --- |
|  |  |  | with dementia, their caregivers and their relatives and being responsive to continually changing needs and abilities of the resident.(28) |
| Cultural impact | Providing culturally appropriate care is particularly important for people with dementia. | 2(28, 42) | Ways to provide culturally appropriate dementia care included:<br>- acknowledging the individuality of residents, catering to their needs and choices, and connecting them with familiar rituals and activities(28)<br>- speaking the language of the resident in order to maintain familiarity and connectedness(42)<br>- having a diverse workforce with multilingual staff to support residents with one-on-one care (28, 42)<br>- placing residents of a similar cultural background in proximity within the home(28)<br>- staff having an understanding of culturally and linguistically diverse backgrounds(28)<br>- staff actively seeking to collaborate with cultural communities to assist in staff training, to organising cultural-specific activities and to provide culturally appropriate meals.(28, 42) |
| Care planning | Care planning is a fundamental aspect of personalised dementia care practice | 3(17, 29, 39) | 1. Care planning must be individualised, designed to include the voice of residents and their family members and must reflect the residents' history, needs and wishes.(17)<br>2. Basic needs such as identity, comfort, inclusion, attachment, and occupation, are particularly important to comprehensively document.(29)<br>3. The following areas should also be identified and provided for residents in care plans:<br>- any spiritual preferences and practices and how these needs are met<br>- details of what items or people the resident has formed attachments to<br>- details of what is psychologically comforting for the resident<br>- important community links and how staff can facilitate these<br>- details of how to maintain and support the resident's unique identity<br>- details of how the resident wishes to be included in communal activities<br>- if relevant, details on a lasting power of attorney or enduring power of attorney.(39)<br>4. Failure to comprehensively record information on psychosocial aspects can hinder understanding whether important basic needs have been considered or whether interventions are appropriate.(29) |
| Meaningful engagement | Meaningful engagement is a critical dimension of quality of life and quality of care for persons living with dementia. | 2(19, 20) | 1. Staff caring for residents with advanced dementia relied on being sensitive to non-verbal signs such as emotional expression, touch, and posture to know when and how to attempt meaningful connections.<br>2. Staff meaningfully engaged with residents by getting to know the resident, learning their communication patterns, speaking clearly using short sentences, making eye contact, giving resident time to respond.(19)<br>3. Other approaches included approaching residents in a kind, gentle and caring way at a pace that suits the resident; greeting |

| Theme | Description | Studies addressing theme | Key findings |
| --- | --- | --- | --- |
|  |  |  | a person by name or using nicknames; staff members introducing themselves, using humour, sharing information about their own hobbies or family are part of meaningful engagement and staff exercising patience and perseverance.(19, 20)<br>Staff felt constrained in forming connections by having other tasks to do.(19)<br>It must be emphasised to staff that meaningful engagement is a core part of their role.(19)<br>All encounters, even those that are routine e.g. personal care, should be viewed as having potential for meaningful engagement. Encounters could also include ones with non-clinical personnel.(19, 20) |
| Eating and meals | Mealtimes are important in the care of people with dementia. | 3(30, 32, 46) | Mealtimes were found to be task based with little engagement with and among residents.(46)<br>Practice changes included:<br>- ensuring greater staff to resident ratios at meal times<br>- increased assistance with meals including monitoring, encouraging, and physically assistance<br>- making changes to the physical environment such as replacing televisions with soft background music, placing condiments on tables for residents to choose, setting the tables and providing adequate light levels during mealtimes.(32)<br>Changes can lead to significant improvement in resident-to-resident interactions, staff interacting with affection to residents, increased communication and connection between health care assistants and family members, and a reduction in task-focused interactions.(30, 32)<br>Practice changes benefitted resident's well-being, led to residents experiencing less pain, consuming at least the minimum recommended fluids, and being more at ease and engaged with staff.(30) |
| The role of family members | Involving family members is key in providing good care for people with dementia in nursing homes. | 5(14, 15, 17, 31, 47) | Effective communication between staff and family members has been reported to be inadequate, in particular in relation to information exchange and shared decision making.(17, 31)<br>Family members reported an absence of friendly, casual communication with the staff, and sometimes feeling unwelcome. When their relatives died, some family carers felt that there was little consideration of their grief.(15)<br>Insufficient support for family members in relation to discussions about end of life care has been shown to lead to subsequent resistance to palliative care.(31)<br>Family members have a unique contribution to make to person-centred assessment, including capturing and sharing life history to identify unique elements of the person.(14, 31, 35)<br>When family members are included, this leads to a person-centred approach, contributes to residents' well-being and supports connection of the residents with their community beyond the facility.(15) |
| Restrictive practices | Restrictive practices have a negative impact | 3(26, 30, 38) | Physical restraint use is a potential quality indicator for dementia care nursing homes.(30, 38)<br>Physical restraint has been shown to be unequivocally damaging |

| Theme | Description | Studies addressing theme | Key findings |
| --- | --- | --- | --- |
|  | on caring for people with dementia in nursing homes. |  | for residents, who experienced more pain and depressive symptoms, lower BMIs, lower scores on a standardised quality of life measure, and they were less at ease and less engaged with staff.(30)<br>3. The most common restraint used on people with dementia was restricted movement through not providing mobility aids,(26) or residents with dementia not being included in the full range of external activities on offer to other residents due to staff perception that public spaces outside of the nursing homes were not physically accessible.(26) |
| Psychotropic medications | This theme discusses practices of administering psychotropic medications to people with dementia in nursing homes. | 2(24, 38) | 1. Staff are aware of their need to keep the use of anti-psychotic drug prescribing to a minimum however barriers include concerns about the physical safety of nursing staff and residents.(24, 38)<br>2. Recommendation to reduce the use of psychotropic drugs included ruling out medical causes of difficult behaviours, trying nonpharmacological alternatives first (for example, music, pet therapy or physical activity), as well as non-antipsychotic drugs, such as mood stabilizers.(24)<br>3. Residents with dementia may also have additional mental illness that necessitates the prescription of anti-psychotics.(24, 38)<br>4. Individualised, person-centred care approaches were deemed essential to reducing use of anti-psychotics.(38) |
| Activities | Meaningful activities and stimulation are important for residents with dementia. | 6(15, 18, 25, 27, 28, 34) | 1. Residents expressed joy or anticipation when invited to events and activities, or when they received visits from children and others in the local community.(27)<br>2. Gaps in participation in everyday occupations among persons with dementia included:<br>- residents "wanting to do but not doing" activities such as minor shopping, hobbies, outdoor activities<br>- residents "doing but did not want to do" activities such as listening to the radio or watching TV.(34)<br>3. Residents with advanced dementia were often not able to benefit from the activities and possibilities provided in the nursing home, since this required additional support.(25)<br>4. It is possible to find meaningful, stimulating, and person-tailored activities for all residents through securing valuable information from the resident or family members.(15, 18, 25, 28)<br>5. For activities to be meaningful and person-tailored:<br>- staff must work with the resident, using creativity, sensitivity, and a try-and-fail approach(18)<br>- staff must adapt activities to the resident's current condition<br>- additional staff and volunteers may be required.(25) |
| Materials | Due regard for material belongings to people with dementia in nursing homes is necessary. | 1(21) | 1. Residents' interactions with their material surroundings provides them with the ability to continue routine practices such as organising their surroundings, cleaning and hosting.<br>2. Residents are often not involved in deciding which belongings they wanted to take to a nursing home.<br>3. Residents depend upon individual staff members, as to whether they can access a particular object, rather than through an organised and structured care planning and risk management |

| Theme | Description | Studies addressing theme | Key findings |
| --- | --- | --- | --- |
|  |  |  | process.<br>4. Guidance is needed in the management of possessions, and an understanding of the importance of object–person relations for people with dementia, in policy and practice documentation.(21) |
| Health Care | A person-centred approach to healthcare is needed for people with dementia in nursing homes. | 4(14, 23, 31, 40) | 1. Nursing homes must go beyond providing basic health care to also address residents’ emotional, social and practical needs.(40)<br>2. An emphasis should be placed on social and emotional bonding, and less on a medical approach.(31)<br>3. A disease-focused perspective to health care should be replaced with a person-centred one,(14) which allows the negotiation of health care based on what matters to residents.(23, 40)<br>4. Healthcare providers should give residents a voice, and include specific professional approaches to sustain personhood, build relationships and support residents to engage in activities.(23)<br>5. A person-centred approach to health care is challenged in nursing homes with the coexistence of disability, cognitive decline, and chronic conditions.(40)<br>6. Healthcare providers may not have the necessary time resources to deal with the key aspects of the patient experience because they are affected by wider organisational systems.<br>7. Achieving health-care quality in nursing homes requires finding a balance between the general routine and the residents’ individual preferences and needs.(23) |
| End of life care | There are both barriers and potential for better outcomes for people with dementia receiving end of life care in nursing homes. | 3(31, 33, 36) | 1. Barriers included:<br>- a lack of knowledge and awareness among nursing home staff which resulted in staff and family not seeing the need for palliative care causing a delay in addressing care needs<br>- care goals or treatment plans not documented properly<br>- a poor interdisciplinary team approach<br>- a lack of continuous care due to staff turnover, poor information transfer and collaboration.(31)<br>2. End-of-life care outcomes were reported to be better in nursing homes compared to hospitals.(36)<br>3. Advanced care planning and individual assessments are needed to avoid referral to hospitals.(33, 36) |
| Staffing | Sufficient staffing levels and consistency in staff is crucial in providing care for people with dementia. | 7(16, 17, 24, 31, 33, 42, 45) | 1. A lack of resources in nursing homes negatively impacts the care provided to residents living with dementia(31, 33, 45) and had the following impact:<br>- High workloads, clinical inexperience, poor leadership, priority given to cost containment, and the lack of dementia-specific staff training(16)<br>- Time restrictions on staff’s interactions with the residents with the focus on routine tasks.(17)<br>- Over-extended staff leading to missing key indicators (i.e. deconditioning) due to apathy or feeling overworked(33), and in inappropriate prescription of antipsychotics.(24)<br>- Challenges in providing care to those from culturally and linguistically diverse backgrounds.(42)<br>2. Consistent dementia care is hindered by high rotation of staff(17) and use of interim and temporary staff for administrative and clinical positions.(33) |

| Theme | Description | Studies addressing theme | Key findings |
| --- | --- | --- | --- |
|  |  |  | 3. Increased staff resources with an emphasis on the staff retention, recruitment and resource adequacy are needed to improve the continuity of care.(17, 31) |
| Staff training | This theme highlights the importance of dementia specific staff training. | 8(14, 16, 17, 20, 24, 31, 41, 42) | There is a need for increased dementia-specific staff training on:<br>- the provision of palliative dementia care including symptom recognition, using standardised instruments, communication with families and advanced care planning(31)<br>- managing behaviours and the use of antipsychotic drugs(24)<br>- applying specific person-centred approaches (14, 20, 41)<br>- culturally appropriate forms of respect (24)<br>- communication with residents.(42) |

### Vignettes

#### Provider

This vignette provides examples of quality dementia care from the provider’s point of view.

It addresses a number of themes identified in this systematic review (table 1) and illustrates how these can be implemented into practice. The themes addressed in this vignette include: person-centred care,(14, 28, 35, 40, 41, 44) cultural impact,(28, 42) meaningful engagement,(19, 20) eating and meals,(30, 32) the role of family members,(14, 17, 30, 31, 35, 42) restrictive practices,(25, 26) psychotropic medication,(25, 26) materials,(21) end of life care(31, 33, 36) and staffing.(16, 17, 31, 33, 45) Niamh is the registered provider of a 30-bed purpose-built nursing home facility. The facility provides 24 hour nursing care to residents for continuing, palliative and dementia care. Most residents are 65 years or older.

Niamh recognises the importance of person-centred care and views it as a practical and manageable approach for providing meaningful care to residents in the centre. She has seen its positive impact on quality of care, resident well-being, and staff satisfaction and retention. She promotes it as a culture not as an “add on” to something to “buy into” but something that infiltrates every activity within the nursing home. She encourages staff members to demonstrate this in the language they use and in the mentoring they provide. She instils practices such as placing a focus on behavioural support, promoting person-centeredness above protocols, insuring training on person-centred culture, shifting mind-sets from generalised to personalised care, valuing implementation flexibility, and being responsive to continually changing needs and abilities of the residents.

Niamh has hired a workforce with a varied demographic profile to support residents from diverse backgrounds. Where possible she ensures residents of similar cultural backgrounds are placed in proximity within the nursing home. She ensures staff have an understanding of residents from culturally and linguistically diverse backgrounds through providing training. Niamh organises cultural-specific activities and provides culturally appropriate meals to residents of different backgrounds, where requested.

Through having appropriate staff-to-resident ratios, Niamh encourages and facilitates her staff to form connections with residents and to meaningfully engage with them. She encourages a culture where all encounters, even those that are routine, are viewed as having potential for meaningful engagement. These encounters could also include ones with non-activity personnel, including kitchen, housekeeping, other staff, and visitors.

Niamh recognises how crucial meals and mealtime practices are to resident’s well-being and employs greater staff-to-resident ratios at meal times to support monitoring, encouraging, and physically assisting with meals. She has experienced that when these practices are in place, residents were more likely to consume food and drink, and are more at ease and engaged with the staff members.

Niamh is welcoming to family members visiting the nursing home. She has a friendly rapport with family members and sees them as being integral to developing a sense of connection with the residents within the facility. She knows how high involvement of family members in their relatives care contributes to resident wellbeing and their care quality. She acknowledges family members as equal partners in providing care and encourages them to be take part in all aspects of the resident’s daily life.

Niamh is aware of the impact physical restraint use has on care quality. She also acknowledges the challenges faced by staff to facilitate resident’s movements. She promotes their freedom of movement through adequate staffing and the provision of mobility aids. She ensures all residents are able to take advantage and benefit from the activities and possibilities provided in the nursing home even if it requires additional staff and volunteers.

Niamh is aware of the need to keep anti-psychotic drug prescribing to a minimum; she experiences barriers to reducing anti-psychotic drugs use as some staff members express concerns about their own physical safety without these medications. She regularly facilitates staff training on managing behaviours and use of antipsychotic drugs. She also acknowledges that some residents with dementia may have additional mental illness that necessitates the prescription of anti-psychotics. For example, patients with dementia might have bipolar that warrants antipsychotic use.

Niamh understands the importance of a resident’s personal possessions to them. She includes object–person relations in the nursing home’s policy and practice documentation; this explains to staff the value in a person continuing to use their own possessions as this supports connectivity to themselves and their life before the nursing home.

Many of the residents in the centre require palliative care. Niamh enables processes for effective information transfer and effective collaboration with healthcare professionals outside of the facility. Through advanced care planning and individual assessments, the majority of residents are managed in the facility until the end of their lives. She facilitates training in the provision of palliative dementia care, in particular in symptom recognition, using standardised instruments, communication with families and advanced care planning.

Niamh has experienced the impact a lack of staffing resources has on residents living with dementia. This has included the loss of administrative staff and use of interim and temporary staff for administrative and clinical positions. This has led to high workloads and clinical inexperience among staff, and the lack of support and dementia-specific staff training. She recognises this as poor leadership and priority given to cost containment above care delivery. Niamh ensures she has the required staffing to improve the continuity of care, with a balance between organisational and administrative routines and the resident’s needs.

#### Staff

This vignette provides examples of quality dementia care from a staff member’s point of view. It covers the following themes: determinant of care quality,(13, 16, 39) person-centred care,(13, 16, 39) cultural impact,(28, 42) meaningful engagement,(19, 20) eating and meals,(30, 32) the role of family members,(14, 15, 17, 30, 31, 35, 42) care planning,(28, 29) activities,(15, 18, 25, 28) end of life care,(31) and staffing.(16, 17, 28, 32, 33)

Sarah is a nurse working in a nursing home registered to accommodate a maximum of 35 residents. She works to provide comfort to each resident she meets and triesto reach the person within the disease. She provides “person-centred dementia care” which is about forming connections and relationships for well-being. She uses techniques to meaningfully engage with residents which include speaking clearly using short sentences, making eye contact, and giving the resident time to respond. She relies on being sensitive to non-verbal signs in the resident such as emotional expression, touch, and posture when attempting meaningful connections. She approaches residents in a kind, gentle and caring way at a pace that suits them. She greets residents by their name or uses their nicknames; she also gives their own name, uses humour and shares information about her own life. She recognises that patience and perseverance are key to engaging.

Sarah spends time getting to know the residents and recognises this as a core part of her role. She sees all encounters, even routine encounters, such as personal care, as having potential for meaningful engagement. During mealtimes, she sees the importance of providing the right atmosphere which includes playing soft background music, placing condiments on tables for residents to choose, setting the tables and providing adequate light levels. Where family are present she involves them in creating and implementing these person-centred mealtime practices.

Sarah works closely with families of residents, having frequent conversations, and informing them about everyday changes in their relatives’ care needs. She lets them know they can ask her if they had queries. She is friendly and engages in casual communication, and ensures they feel welcome. She recognises the unique contribution they make to person-centred assessment, including capturing and sharing life history to identify unique elements of the person.

Sarah has received training to be aware and understand the need for a palliative approach for people with dementia and to avoid delay in addressing their care needs. She provides high-quality end of life care through updating and documenting care goals or treatment plans, sharing information correctly, and working closely and collaborating with the interdisciplinary team. When their relatives have died, she considers the family’s grief.

She strives to have an understanding of residents from culturally and linguistically diverse backgrounds, through connecting them with rituals and activities with which they were likely to be familiar. Where residents do not speak the same language, she engages with them through actions such as developing cards or pictures in the residents’ language, using body language for communication, or by translating care plans into the resident’s language.

Sarah understands that increased focus on the perspectives and experiences of the resident in care planning can create an environment that respects and maintains their individuality.

She works with residents to ensure their care planning is individualised and includes the voice of the resident and their family members. She also ensures it reflects the residents’ history, needs and wishes. Sarah integrates each person’s care plans with extensive information on each person’s favourite soothers and stressors. She continually updates it with the resident’s changing needs and abilities.

Sarah is aware of the importance of finding meaningful, stimulating, and person-tailored activities for residents living with dementia. She works to secure valuable information regarding resident’s choices and desires. She also speaks to the resident’s family members to provide information about former meaningful activities. She ensures activities are meaningful by working with the resident in the moment, using creativity, sensitivity, flexibility and a try-and-fail approach. She uses her professional knowledge to adapt activities to the resident’s current condition, as the illness progresses.

Over the years, Sarah has experience a lack of staffing resources within the nursing home which has led to high workloads and a lack of support and dementia-specific staff training. She has experienced time restrictions which have tied her to routine and task based duties, limiting her interactions with residents. Being over-extended has led her to missing key indicators (such as deconditioning and changes occurring in people with dementia) due to apathy and fatigue. Appropriate staffing in the nursing home has supported her to implement a person-centred approach in her daily work.

#### Resident

This vignette provides examples of quality dementia care from a resident’s point of view. It covers the following themes: determinants of quality care (13, 15, 16, 30, 37, 38, 39), outcomes (18, 30, 39), person-centred care (14, 17, 18, 28, 32, 35, 41, 44), cultural impact (28, 42) care planning (17, 29, 39), meaningful engagement (19, 20), eating and meals (30, 32, 46), the role of family members (15, 17, 31), restrictive practices (26, 30, 38), activities (15, 18, 25, 27, 28, 34), materials (21), health care (14, 23, 31, 40), end of life care (31, 33, 36), staffing (16, 17, 24, 31, 33, 42, 45) and staff training (14, 16, 17, 20, 24, 31, 41, 42).

Kanan is 72 years old man who recently became a resident in a nursing home located in a small town in the west of Ireland. He has lived in Ireland for almost twenty years but is originally from Odisha in India. He had been living with his wife Nita until she was unable to meet his care needs. He was diagnosed with dementia one year prior to admission to the nursing home.

Kanan, Nita and the Director of Nursing, Martin have a meeting to discuss how to make Kanan feel at home in the nursing home. Martin asks them about Kanans hobbies, interests, and community ties. Kanan tells Martin that he was a coach at the local badminton club until a couple of years ago. Martin asks Nita if she would like to bring Kanan to club match days on every second Sunday. Nita says that she would be delighted to. Nita asks if she will be allowed to bring in some of Kanans personal belongings such as electric razor, bedside radio, chess board and books. Martin agrees and says residents are encouraged to bring personal materials because they should feel at home in the place where they live.

After the meeting, Martin asks Nita for a private chat. He assures her that Kanan will have an excellent standard of care. He explains that all of the nursing home staff have had dementia specific training, there is a good staff to resident ratio and residents engage in a wide variety of activities.

A typical week for Kanan involves being greeted by a friendly and patient staff member each morning who assists him with getting dressed and administers his medication. Kanan sits with other residents and staff members at mealtimes. Mealtime seating arrangements are regularly changed to facilitate social interaction. Residents and staff share stories and talk about current events. Kanan engages in activities such as playing chess, yoga and outdoor nature sketching. Outdoor spaces are monitored but accessible for Kanan and other residents. Staff are trained to limit the use of restrictive practices and to foster a sense of freedom and homeliness for residents.

After several years, Kanan’s physical condition deteriorated and his doctor advised the Director of Nursing that he may need to develop a plan for end of life care. The director meets with Nita and explains that it’s important, even for people with dementia. Together, with Kanan, they decide he would be happiest to remain in the nursing home as he is comfortable there and has many friends. Nita explains that as part of their Hindu culture she would like Kanan to be visited by a Hindu priest, for Kanan to be placed on the floor on a clean mat as his time of death approaches and, once he has passed that only family members should touch him. The director assures Nita that these requests will be honoured. She calls a meeting with the care staff to explain the importance of Nita’s requests.

#### Family Member

This vignette provides examples of quality dementia care from a family member’s point of view. It covers the following themes: outcomes (18, 30, 39), person-centred care (14, 17, 18, 28, 32, 35, 41, 44), care planning (17, 29, 39), the role of family members (15, 17, 31), activities (15, 18, 25, 27, 28, 34) and materials (21).

Harriet’s mother Mary is a nursing home resident living with dementia. Mary has been a nursing home resident for six months and received a dementia diagnosis two months ago. Harriet receives a phone call from a member of staff at the nursing home asking her to come in to talk about Mary and about ways to make her mother feel at home.

Harriet meets with Susan, the Director of Nursing. The director has a friendly and casual demeanour which puts Harriet at ease. She is offered tea and invited to sit in a comfortable, area of the nursing home. The director explains that it would be beneficial to Mary if Harriet could provide some information and assistance regarding a number of aspects related to her mother’s care.

Harriet is told that care planning is a fundamental aspect of personalised dementia care and that her input would be hugely beneficial in developing a high quality, person centred care plan for Mary. She is asked if Mary has any community links. Harriet tells the director that Mary has been part of a local book club for many years. Susan asks if Harriet could reach out to the book club with a view to having occasional meetings at the nursing home as a way to keep Mary involved in the club.

Harriet is then asked about Mary’s hobbies and interests. Mary has always had a keen interest in gardening and horticulture. Susan explains that this is a common hobby among residents and as a result she is having discussions with the senior manager about procuring a polytunnel and outdoor space so that residents can continue to actively engage with this interest. She explains that they are also looking to develop a volunteer scheme with a local secondary school so that students could help residents with physically demanding tasks related to this.

Finally, Susan tells Harriet that it is important for family members to be involved in decision making and asked her if she would like to be contacted regularly when decisions need to be made in relation to Mary’s care. Harriet agreed.

After the meeting, Harriet felt a sense of comfort and connection in relation to her mother’s care and her own involvement. She also felt that her mother’s quality of life needs were important to the staff at the nursing home.

## Discussion

In recent years, a number of reports have been published demonstrating the growing prevalence of dementia along with an aging population. These reports provide up-to-date knowledge on the prevention, intervention and care of people dementia.(7, 8, 9) While these reports provide overarching recommendations on intervention and care, they do not focus on identifying the characteristics of quality in the provision of care for people living with dementia in nursing homes settings. This review advances knowledge in this area though the identification of sixteen good practice themes. The themes were: determinants of care quality, outcomes, person-centred care, cultural impact, care planning, meaningful engagement, eating and meals, the role of family members, restrictive practices, psychotropic medications, activities, materials, health care, end of life care, staffing and staff training.

The findings from this review promote providing a person-centred approach to dementia care in nursing homes where residents are involved in all decisions around their care and support and staff work closely with both the resident living with dementia, their caregivers and their relatives. The emphasis must be placed on effective communication with resident and their family, and recognising the importance of connections and relationships on the well-being of residents with dementia.(28) A person-centred approach must infiltrate all activities conducted in a nursing home including communication, assessment,(14, 31, 35) provision of meals,(32) and the provision of medical treatment. Training in specific person-centred approaches is essential to support improved quality-of-life for people living with dementia.(14, 20, 41)

Effective communication with the resident with dementia is a key component of high quality care. Emphasising to staff the importance of getting to know residents, learning their communication patterns, and what techniques staff can use to effectively communicate with the resident is key to meaningful engagement.(15, 17, 31) It is also important for staff to know that effective communication is a key part of their role and any encounter from all staff members should be seen as an opportunity to meaningfully engage with the resident.(19, 20) Where the resident does not speak the same language as staff, staff must use other methods to ensure continued communication with the resident, for example through body language or pictures.(28, 42) Effective communication from staff extends beyond the resident to the resident’s family and or caregivers. Consideration for family member’s experiences, shared decision-making between staff and family members, increase communication, and formal processes for sharing important information is needed. Staff must also recognise the importance of and support communication between the resident and their family members to ensure connection with their community beyond the nursing home.(14, 15, 17, 31, 35, 42)

The findings highlight how people living with dementia were often denied their right to freedom, autonomy and participation in society. This was often attributed to a lack of resources, a lack of individualised care, or it was suggested as being an unquestioned aspect of care home culture.(26, 30, 38) It included residents with dementia being denied access to a full range of external activities on offer to other residents or being subjected to activities they did not want to do;(25, 34) residents not being involved in deciding which belongings they wanted to take to a nursing home or not being able to access their belongings;(21, 25) residents being isolated from their families and or communities,(17, 28, 30, 35, 42) and residents being physically or chemically restrained.(24, 26, 30, 38) Additional resources, resident participation, structured care planning, risk management processes and integrating residents into the community are essential for overcoming these barriers.(21, 24)

Challenges in providing high quality care for residents with dementia in nursing homes included staff shortages which often forced staff to prioritise task-based duties,(28) High work-loads also impacted meaningful engagement with many staff feeling constrained by having other tasks to do and having other residents to attend to.(19, 20) Staff also experienced challenges associated with shifting mind-sets from generalised to personalised care, which in practice disrupted social norms for nursing home staff.(17, 32) Healthcare providers were also challenged by not having the necessary time resources to deal with the key aspects of the patient experience because they are affected by wider organisational systems.(23) Furthermore, there was a lack of support and dementia-specific staff training prevalent within nursing homes with training required to enhance awareness of person-centred care,(14, 20, 41) palliative dementia care,(24, 31) managing behaviours and use of anti-psychotropic drugs,(24) complex care needs of dementia(16, 17) and culturally appropriate care.(42)

### Strengths and Limitations

This review has limitations. We only included published peer-reviewed literature. It is possible that many more articles reporting on the characteristics of quality in the provision of dementia care in nursing homes through grey literature. The wide heterogeneity of included studies (in terms of study designs, key areas of best practice addressed, and samples included in individual studies) can also be considered a limitation, as this only enabled us to make broad generalisations without consideration individual study quality. A review protocol was prepared but not published. Notwithstanding these limitations, this review is important in drawing attention to the characteristics of quality in the provision of dementia care in nursing homes based on the most up-to-date research. To the authors’ knowledge, this is the first published review in the last decade to summarise and synthesise evidence on what constitutes high quality care specifically in the nursing home sector. It also provides insights that may be of use to those delivering care in nursing homes but also to policymakers and researchers when conducting future work in this field.

### Conclusion

This review summarises and synthesises evidence on what constitutes high quality dementia care in nursing homes. Sixteen themes were identified and can be used as the basis for structuring quality improvement interventions in relation to people living with dementia. They findings also provide valuable insights which will be of use to those delivering care in nursing homes and to regulators, policymakers and researchers when conducting future work in this field.

## Data Availability

All relevant data are within the manuscript and its Supporting Information files.

