## Supplemental file for "Good practice in the provision of care for people living with dementia in nursing homes: a systematic review"

Supplementary file: Search Terms, good practice in the provision of care for people living with dementia in nursing homes: a systematic review

The search terms for one electronic database (MEDLINE) are provided below. The other databases were searched using a similar approach.

Search terms used for the electronic database MEDLINE for the review of good practice in the provision of care for people with dementia

| # | Search terms |
| --- | --- |
| #1 | (MH "Aged") |
| #2 | TI AB ("older adult*" OR "older person*" OR “older people" OR senior* OR aged OR elderly OR geriatric* OR citizen* OR resident* OR “service user*”) |
| #3 | ("Residential Facilities"[Mesh]) OR ("Nursing Homes"[Mesh]) OR ("Intermediate Care Facilities"[Mesh]) OR ("Skilled Nursing Facilities"[Mesh]) OR ("Assisted Living Facilities"[Mesh]) OR ("Homes for the Aged"[Mesh]) OR ("Group (Homes"[Mesh]) OR ("Long-Term Care"[Mesh]) |
| #4 | TI AB ((residential OR institution*) N2 (care OR home* OR facilit* OR center* OR centre*)) OR ((long-term OR longterm OR long term) N2 (care OR service*)) OR (retirement N1 (home* OR village* OR communit*)) OR “nursing home*” OR “skilled nursing facilit*” OR “care home*” OR “aged care facilit*” OR “old age home” OR “rest home*” OR “service home*” OR “sheltered hous*” OR ((assisted OR supported) N1 living) OR “hous* for the elderly” OR “home* for the aged” |
| #5 | ("Quality Assurance, Health Care"[Mesh]) OR ("Quality Indicators, Health Care"[Mesh]) OR ("Program Evaluation"[Mesh]) OR ("Facility Design and Construction"[Mesh]) OR ("Health Facilities, Proprietary"[Mesh]) OR ("Health Facility Closure"[Mesh]) OR ("Health Facility Merger"[Mesh]) OR ("Health Facility Size"[Mesh]) OR ("Outcome and Process Assessment, Health Care"[Mesh]) OR ("Patient Care"[Mesh]) OR ("Patient Satisfaction"[Mesh]) OR ("Public Reporting of Healthcare Data"[Mesh]) |
| #6 | TI AB (quality OR infrastructure OR market* OR sector* OR organisation* OR organisation* OR staff* OR personnel OR workforce OR recruit* OR retention OR “care plan*” OR “care need*” OR “behaviour support” OR “behavior support” OR restraint* OR “restrictive practice*” OR “infection prevention and control” OR “hand hygiene” OR “pain management” OR prescri* OR  “care outcome*” OR hospitalisation* OR hospitalization* OR admission* OR discharge* OR fall* OR "functional ability" OR "functional capacity" OR "functional decline" OR "activities of daily living” OR “pressure ulcer*” OR “pressure sore*” OR “pressure injur*”) |
| #7 | MH "Dementia") OR (MH "Dementia, Vascular") OR (MH "Lewy Body Disease") OR (MH "Mixed Dementias") OR (MH "Alzheimer Disease") OR (MH "Dementia, Multi-Infarct") |
| #8 | TI ( (Dementia OR Alzheimer OR “vascular dementia”) ) OR AB ( (Dementia OR Alzheimer OR “vascular dementia”) ) |

Note: Last searched on 06/034/2023. #2, #4, and #6 searched for Title OR Abstract fields only. Filters applied: Language = English; Years = 2020-2023.
