## Supplemental Table 1 for "Good practice in the provision of care for people living with dementia in nursing homes: a systematic review"

**Supplementary table 1: Characteristics of included studies**

| **Author**  **(year)** | **Jurisdiction** | **Study**  **design** | **Aim** | **Characteristics**  **of**  **sample** | **Overview of Results** | **Criteria or elements**  **of what constitutes high quality of care** |
| --- | --- | --- | --- | --- | --- | --- |
| Anderson and Blair (2020) | Australia | Cross-sectional study | To examine associations between quality of care, and broadly defined quality of life for residents with dementia. It also aims to establish where it is most useful to intervene in quality of care in order to increase quality of life for people with dementia. | Older adults in residential care with dementia (n = 247), their families/care partners (n = 225), managers (n = 12) and staff (n = 235) of 12 residential aged care facilities. | - Quality of care (QOC) (determined by staff treatment, assistance at meals, physical restraint, psychotropic medications, adverse incidents, person-centred care scale score and quality of care scale score) had an immediate impact on resident’s pain, depression, QOL scale score, Body Mass Index, ease/engagement with staff, and food and fluid intake. - Restraint was a predictor of poor outcomes for residents. - QOC variables significantly predicted pain reduction. - Restraint use had an immediate effect on depression. - Ease and engagement were higher where more assistance was provided with meals. - QOC variables had an immediate effect on food and fluid intake. | - This study demonstrated that what staff do with the residents and the way they do it has an effects on the QOC of residents with dementia in nursing home. - Physical restraint is unequivocally damaging for residents who experienced more pain, depressive symptoms, ate less and had lower BMIs, had lower quality of life measure, and they were less engaged. - Providing greater assistance led to residents experiencing less pain, greater consumption at least the minimum recommended fluids, and are more engaged with staff with greater staff to resident ratios at meal times. - Increased psychotropic medication use resulted in reduced food intake, both immediately and longer-term. - Adverse incidents had no effect, suggesting it is more the day-to-day integrated interactions between staff and residents that is important rather than activities. |
| Balqis et al. 2020 | Indonesia | Qualitative study | To find out the true support needed by people with dementia while living in nursing homes. | Ten elderly participants with mild/moderate dementia. | - People with dementia need holistic support including (a) biological support which constitutes as food needs, drinking needs, and bathing needs; (2) psychological support through the spirit of life, compassion and optimism; (3) social support which is the desire to remain socialised even though the elderly experienced declines; (4) spiritual support to help get closer to God, helps in worship, help during serenity, and help to have peacefully end of life. | - Biological supports were often considered a daily routine without regard to the quality of the fulfilment of these services. - In term of social supports, these include the need to have regular and meaningful conversations with people around them. In line with other studies, social support is needed by the elderly with dementia to provide mutual support, create a sense of belonging and provide information needed. - People with dementia wish to be actively involved in the selection of care in the future, for dementia and in end of life care. |
| Bavelaar et al. (2021) | The Netherlands | Cross-sectional study | To address what are the barriers to providing high-quality palliative care in dementia according to elderly care physicians. | Dutch registered elderly care physicians. | The barriers to providing good quality palliative care in dementia include:   - Beliefs and lack of knowledge, awareness or understanding. - Families, hospital doctors, nursing staff, and the public did not see the need for a palliative approach for people with dementia. - Obstacles in recognising and addressing care needs i.e. managing decline, discomfort, and starting the palliative phase. - Poor interdisciplinary team approach and consensus. Care was not continuous because of high staff turnover, poor information transfer, and poor collaboration between healthcare professionals. - Palliative care terminology was used inconsistently, and uncertainty remained about what a palliative care approach entailed. - Limited use or availability or resources such as a lack of time and poor staffing negatively impacted the care provided. - Poor family support and involvement. Frequently family did not feel ready to part with their relative-hence resisting palliative care. Underlying this resistance was insufficient support for families, as nursing staff were not able to timely discuss the end of life. | - Nursing home staff need to increase the frequency of their conversations with family and provide counselling. Nursing home staff should engage family in daily care tasks and improve their interaction with cultures. - Multidisciplinary training could enhance palliative care knowledge and overcome several barriers related to limited staff resources. - More staff and decreased (administrative) workload would reduce lack of time. Additionally, investing in having the same healthcare professional attending the same patient and family would facilitate relationships. - Public education on palliative care could help address barriers to providing high-quality palliative care in dementia such as the perceived unrealistic public image of prolonging or ending life, the denial of dementia diagnosis or prognosis by some families, and the difficulties in recognising and addressing care needs. |
| Bergstrom et al. (2023) | Sweden | Cross-sectional study | To describe gaps in participation in everyday occupations among people with dementia living in nursing homes. | Relatives of residents with dementia (n=98) | - The most common occupational gap was “wanting to do but did not do” and was found in shopping, hobbies, travelling for pleasure, outdoor activities, and playing the lottery, crossword puzzles, and board games. Another was “did but did not want to do” was in listening to the radio, watching TV/videos. - The items that the respondents reported that the persons in the target group participated in and wanted to do were visiting with partner/children, listening to radio/watching TV, and visiting with relatives and friends. The items that the respondents reported that - They did not participate in and did not want to do were using computers, involvement in societies, clubs, or unions and practicing religion. - Persons with dementia are often dependent on another person’s assistance to different degrees. The realisation of not having a possibility to participate could possibly impact the actual desire to participate and thus a response of not wanting to do. | - Realising the importance of nursing home residents’ participation in occupations is fundamental, but the first step in achieving participation in securing valuable information regarding individuals’ choices and desires. - In this study, proxies to people with dementia seemed to be an appropriate source of information since they most likely have extensive knowledge of the person before as well as after being admitted to the nursing home. - After obtaining information regarding the desires for participation, a possible solution for increasing participation may involve collaboration with the respondents, other interested acquaintances, or close family members. Occupational therapists could then contribute with specific knowledge regarding enabling occupations as well as empowering family members to assist with participate in desired occupations. |
| Boumans et al. (2021) | The Netherlands | Cross-sectional study | To explore the association between staff characteristics (age, education level, years of work experience and function, i.e., care or welfare) and staff attitudes toward perceived person-centered care provision and including informal caregivers in the caregiving process in residential care facilities. | 68 care staff (nurses, nurse assistants, activity counsellors, hostesses, living room caretakers) of two residential care facilities. | - Care staff had a significantly more positive attitude toward informal care provision compared to welfare staff. - Findings showed that in a physical environment that met most of the conditions to realize person-centred care (PCC), both care and welfare staff were convinced that they provide PCC to a relatively high degree. Also, staff members held a rather positive attitude regarding informal care provision. - When comparing care and welfare staff, care staff held a more positive attitude toward informal care provision than welfare staff. Furthermore, a higher age of staff was related to more negative attitudes about their perceived PCC provision. The results could not confirm the existing research that care staff with higher levels of education have more positive attitudes towards residents with dementia. PCC did not differ by years of work experience in long-term care. A reason could be that recently graduated care staff have received more education about PCC than care staff who graduated longer ago. | - PCC provision in residential care facilities does not depend on paid staff only. Forming and maintaining good relationships between staff members, clients, and their families is essential to PCC. - Staff members with a positive attitude toward PCC and the inclusion of family members in care could lead to more PCC for persons with dementia, which contributes to residents’ well-being. - The facility culture of a dementia care facility can evolve over time and administrators of dementia care facilities may be able to adequately target the reasons for the negative attitudes of staff members regarding PCC and informal care by implementing interventions that eliminate or reduce these negative attitudes. |
| Burgess et al. (2023) | USA | Cross-sectional study | To convene a modified Delphi panel to identify potential additional quality indicators (QIs) for nursing home residents with dementia, to generate more QIs to better capture the quality of dementia care. | Ten experts on dementia for Veterans, representing seven medical centers and two national offices. | - Two QIs reached relevance on all three domains: 1) percent of patients with dementia prescribed anti-psychotics (AP) and 2) percent of patients with dementia prescribed benzodiazepines. Percent of CLC patients with dementia with physical restraint use, also reached consensus. - Three QIs reached consensus at the highest median score (9) for importance: 1) percent of patients with dementia with any physical restraint use, 2) AP use, and 3) positive behavioural symptom score. - Four QIs reached consensus at a median score of 7–8 for importance: 1) percent of patients with dementia with any benzodiazepine use, 2) anxiolytic/sedative/hypnotic use, 3) opioid use, and 4) inpatient hospitalization. | - Besides the existing AP QI, consensus was reached for one additional QI: percent of residents with dementia with benzodiazepine use. As benzodiazepines are frequently used for treatment of behavioural disturbances in dementia and carry a similar level of potential harm as APs, results showed that assessing benzodiazepine prescribing alongside AP when evaluating dementia care quality could prove important, useful, and feasible. - There is a need for additional population exclusions from denominators for medication-based QIs. The “right” amount of medication use for nursing home residents with dementia is not zero for any QIs; therefore, as with the AP measure, implementing any of the QI alternatives without relevant denominator exclusions would reduce potentially appropriate medication use. Patients with dementia might have bipolar that warrants AP use, or pain that warrants opioid use. Given the ample evidence for harms with use of benzodiazepines in dementia, policymakers should consider this QI in addition to AP use. However future initiatives aimed at improving the quality of care for nursing home residents with dementia should focus on how to measure the use of recommended evidence-based non-pharmacologic alternatives. |
| Caspar et al. (2021) | Canada | Cross-sectional study | To evaluate the effectiveness of a stakeholder engagement practice change initiative aimed at increasing the provision of person-centred mealtimes in nursing homes. | A nursing home providing care to 12 residents, all of whom were male and had a diagnosis of dementia. | - Statistically significant improvements were noted in all mealtime environment scales. Scales include: physical environment, social environment, relationship and person-centred scale, and overall environment scale. In fact, mealtime environment scores started increasing immediately following the intervention. - Assessment of the physical environment includes elements such as noise levels, seating arrangements, and sufficiency of lighting, aroma of food, decorations and ambiance, and availability of condiments for residents to choose from. Almost all elements of the physical environment showed significant improvement as a result of the intervention. One component of the physical environment that did not show improvement was food aroma. - Resident-to-resident interactions improved over the course of the intervention, as did other positive interactions, such as staff interacting with affection to residents. Task-focused interactions were reduced. - Participants felt the initiative resulted in increased communication and connection between staff and family members. Family felt that they are part of the team. | - This study offers evidence that practice change initiatives that focus on stakeholder engagement can provide a promising method for improving the provision of person-centred mealtime practices at nursing homes. - This included disrupting social norms for RCH, such as being perceived as “lazy” for sitting and spending time with residents during mealtimes. They now advocated that this care practice was “best practice” and essential for PCC. - The study elucidates the importance of cultivating an empowered workforce by implementing practices that enable and encourage collaborative decision making and increase care staffs’ autonomy and self-determination. - Finally, the quality of communication and the level of effective, supportive teamwork were both inextricably intertwined. These organizational factors were also especially salient in their ability to influence the extent to which PCC principles were enacted in practice. - This study’s results indicate that the stakeholder engagement focused practice change initiative is a feasible method for improving PCC mealtime practices in nursing homes. It also adds to the body of knowledge suggesting that improved engagement, teamwork, and collaboration are directly related to the quality of care provided in health care settings. |
| Du Toit et al. (2019) | UK, New Zealand, South Africa and Western Australia | Cross-sectional study | To explore how residential care leadership understand meaningful engagement for residents with dementia from culturally and linguistically diverse (CALD) backgrounds. | One residential facility was included in each country. Participants employed in formal leadership positions at each facility were invited to participate. | - Residents’ choice about whether to participate in activities was the practice likely to be most indicative of PCC. Care plans, which include the life history of residents, is also an invested PCC practice present in the facility. The PCC practice least supported was the quality of the interaction between staff and residents being more important than getting physical tasks done. - In terms of how facilities had successfully engaged residents with dementia from different cultures, examples were (i) involving people; (ii) ways in which language barriers had been overcome; and (iii) specific actions to foster engagement. - All participants valued practical training and staff need to have a manager who is present and practices what they preach. | - Acknowledging resident choice and identities were the two practices regarded as most supportive of PCC in the care facilities. Allowing residents the choice of whether or not they want to participate in everyday activities provides a real sense of freedom - Including life histories in care plans acknowledges residents’ individuality. These practices demand staff time and time demands and staff shortages force staff to prioritize their duties. - The examples of how facilities engaged residents with dementia from different cultures mainly centered on human interaction and addressing language barriers. A wide range of opportunities for human interaction involved people outside the facility including family, volunteers, residents’ cultural and religious communities. - Residents’ engagement could be enhanced by engaging actions, such as developing cards or pictures in the residents’ language, or translating the care plans into residents’ preferred language. The use of body language for communication was perceived to be especially powerful. The use of technology was not suggested. |
| Du Toit et al. (2022) | Australia | Qualitative study | To determine how care practices support the provision of culturally appropriate dementia care for older adults with an Indian heritage living in Australian nursing homes. | Stakeholder groups made up of care staff of Indian heritage, relatives of Indian heritage whose family members are residents in care facilities, and community-dwelling  older adults of Indian heritage. | - Six overlapping themes were found including: (1) embracing a person-centred approach to promote culturally appropriate dementia care, (2) training staff in culturally appropriate forms of respect, (3) the impact of staff ratios on care, (4) the importance of familiarity to meaningful engagement, (5) the importance of food, and (6) the necessity of engaging family and the wider Indian community in residential care activities. - Encapsulated within the theme a person-centred approach, individual’s needs and preferences must be acknowledged, understood, and catered to. - Language and culture barrier was a major obstacle to the provision of culturally appropriate care. The appointment of multilingual staff to support residents with one-on-one care and to act as cultural brokers for sharing important information about the residents with other staff. - Respect for the resident as an elder, respect for gender norms, food preferences, religious rituals, and respect for traditions were a theme. - Staff to resident ratios were noted as one of the biggest challenges to providing good quality care. - Aspects of food and food preparation were associated with health, religious beliefs, enjoyment, and meaningful engagement. Culturally familiar food plays an important role to residents’ heath. | - All stakeholders emphasised the necessity of acknowledging the individuality of the resident, catering to their needs and choices, and connecting them with rituals and activities with which they were likely to be familiar—thereby acknowledging residents’ cultural background. - A diverse workforce that encourages employment of bilingual staff and all staff need an understanding of communication with residents. - Providing opportunities to maintain familiarity by nurturing past passions and interests. - Also noteworthy in our findings was the importance of culturally appropriate food to appropriate dementia care. - Using the Indian community as a resource through involving residents’ relatives and cultural community groups/organisations in cultural activities and celebrations deserves consideration. |
| Fazio et al. (2020) | USA | Qualitative study | To determine the common challenges faced when implementing person-centered, non-pharmacological practices in long-term care and other settings that provide care and programs for people with dementia, and to develop relevant, specific guidance. | A group of 23 administrative leaders and expert providers representing long-term and community-based care settings. | - Heavy workloads, provider turnover, broken equipment, lack of buy-in from key stakeholders, or a failure to maintain regular use of a new practice are common reasons for the abandonment of a practice. - Person-centered practice should not be an “add on” that providers “buy into.” The culture permeates every activity within the organization, from language to mentoring to resource allocation, and envelops every staff member. The foundation of person-centered care begins with leadership. The care recipient and family members are enveloped also, as equal partners. They especially contribute to systemic methods for capturing and sharing life history to identify unique elements of personhood. - Interdisciplinary team processes must be in place to proactively address care recipient needs. - Persons living with dementia commonly experience communication difficulties that interfere with expressing feelings, frustrations, or symptoms. Framed as expressions, the behaviors are clues to the internal state of a person. In person-centered culture, the goal of the care partner is to ask what is this person expressing, what is causing this reaction, and how can one respond to reduce their distress? - Behavioral support can too often become restricted through the “process-oriented” lens of daily care routines, but understanding the concepts of person-centered care must come first in order to recognize how routines may be causing a behavioral expression to occur in the first place. - The methods by which long-term care providers are trained in applying a specific person-centered approach to address dementia-related expressions can themselves reinforce the person-centered values of an organization. - Translating a one-size-fits-all program into person-centered culture might begin with conducting a full review of each person’s favourite soothers and stressors, as gleaned from the person with dementia and their caregivers. Making sure that the person with late stage dementia is completely surrounded by the things that they love the most favourite pillow, blanket, aroma, or musician make the difference in the success of the effort. - The chronic neurodegenerative nature of dementia means that person-centered care must be responsive to continually changing needs and abilities. | - Policy changes are needed that will: - Lead to a reorganization of the way dementia care providers and organizations are incentivized, in order to provide adequate compensation for the time and expertise required to understand, identify, and address behavioral expressions. - Require regular/continuous input from the field, such as through advisory panels, care recipient stakeholder groups, and expert roundtables. This will require funding for behavioural consultation and intervention, and access to the research evidence base. - Encourage greater understanding of person-centered practice in dementia care, including from informal caregivers (family members), formal caregivers, (physicians, nurses, non-medicalstaff members, and administrators) and entities involved in the licensing, monitoring, and regulation of care settings. - It is hoped that the 5 areas of guidance promoted in this articled having a foundational person-centered culture, conceptualizing behaviors as expressions and focusing on behavioral support, identifying antecedents and placing person-centeredness above protocols, insuring training promotes person-centered culture, and valuing implementation flexibility, along with progress in research and policy, will ultimately lead to real provision of effective, person-centered care for behavioral expressions of dementia, regardless of the setting in which care is provided. |
| Fetherstonhaugh et al. (2021) | Australia | Qualitative study | To identify the different factors that make up a well-performing nursing home from the perspectives of family carers. | 21 family members of people with dementia. | - A single overarching global theme was having to be an advocate for their relative in order to have their needs met due to concerns about perceived deficiencies in aspects of care. - The characteristics featuring in a well-performing nursing home related to the following concerns: accountability; communication; staff- resident-family connections; and trust. - Participants described the need for accountability from residential care and staff at all levels. Accountability was comprised of two sub-themes – transparency and individualised care. - Participants felt that the care homes lacked transparency and were unable to care for individual residents’ needs. Participants needed to feel they were included in decision-making, were kept aware of issues, and understood the financial and organisation structures. - An important yet absent aspect of transparency was the ability of the family and older person to meet the staff prior to entry, to gauge its suitability. Many participants described a lack of transparency in their ongoing dealings with nursing homes. - Expectations of individualised care extended to the provision of opportunities for social engagement, in that they were personally meaningful and chosen by their relative. - Open and honest communication with staff and managers was important for carers and poor or no communication fostered a lack of trust, reinforcing the need for advocacy. Participants reported that important information, was not communicated to them and sometimes even actively withheld. Participants emphasised the need for honesty and some reported what they felt were instances of dishonesty to protect the nursing home. - Another thematic category was the need for a sense of connection between the residents and family with the staff. Participants reported that a sense of connection was missing and expressed a desire for the development of relationships between staff, residents and family, in particular so staff could provide companionship to their relative in their absence. Participants wanted their family members to be treated with dignity and respect. Participants also reported that they themselves needed connection with the nursing home; that they felt distant from the staff; and that the absence of friendly communication with the staff meant they sometimes felt unwelcome. When their relatives died, some family carers felt that there was little consideration of their grief. - Another theme was the need for trust. When participants had little or no trust in the staff to provide high quality, safe care, many of them spent more time with their relative. | - Advocacy by family carers of older people living in nursing homes with dementia improved resident care and end-of-life care. - It has been argued that family members are more likely to be advocates because they have extensive knowledge of their relative and therefore notice changes in their health status. - Family carers in this study were most concerned about nursing homes are accountable, both financially and in the provision of care for residents; that there is open communication with residents and the family; that staff-resident-family connections are established and maintained; and that staff can be trusted to deliver the care required by residents. The need for accountability related to the adequacy of staffing numbers and skills as participants perceived this was directly associated with the adequacy of care delivery. - As the carers participating in this study spent a great deal of time at a nursing home, developing a friendly rapport with staff and observing relational interactions between the staff and their relative was thought to be integral to developing this sense of connection. The more that family feel connected to staff and the more they know about them, the more they trust. Staffing skills and numbers are important for building trust. - Overall, these findings demonstrate the importance of positive relationships among family members, staff, and residents. A nursing can only be judged as performing well when what is going on there is transparent, when staff collaborate, communicate and respect family members as care partners, and when family carers can trust staff to deliver individualised care. |
| Gilbert et al. (2021) | Australia | Qualitative study | To understand nurses’ perceptions of quality  nursing care in dementia-specific care units | Nine registered nurses of diverse ages, clinical experience and  cultural backgrounds, working in two separate dementia-specific care units. | - This article reports on the largest theme identified in the study—Labour of Love and the subsequent subthemes: What if that was me, Moments of happiness and the normal everyday things. - Labour of Love identified that quality nursing care was linked to the nurse–patient therapeutic relationship in the dementia-specific care unit. - The awareness of the humanity of the individuals living in the dementia specific care unit. - The importance of the nurse–patient relationship were discussed but felt they were unable to provide more than basic nursing care in the dementia-specific care unit due to high workloads and clinical inexperience. These comments reflected their view of the poor leadership. - The pleasure and satisfaction nurses received when they could reach the person ‘within the disease’ and make a difference to their quality of life was discussed. | - In the absence of a cure for dementia, there is an increased need to recognize the value of the caring and rewarding nurse–patient therapeutic relationship in the dementia-specific care unit. This relationship, characterised by moments of engagement and recognition, has been shown to assist in reducing feelings of social isolation and depression, and can significantly improve the quality of life for people in the dementia-specific care unit. - Understanding what quality nursing care is in dementia-specific care units can provide a coherent picture of the relationships between nurses and individuals with advanced dementia. - The significance of this study lies in its potential to improve the understanding of what constitutes quality nursing care in the dementia specific care unit from the perspectives of nurses. - Findings from this study indicate that nurses provide person-centred care and truly care for people living in the dementia-specific care unit. The provision of person-centred care includes meeting the physical, spiritual and emotional needs of each person, providing comfort for the patient, reaching the person within the disease and making a difference to each person every day. |
| Hamiduzzaman et al. (2020) | Australia | Qualitative study | To explore the factors that shape the dimensions of personalised dementia care in rural nursing homes. | Staff working in five rural aged care homes including clinical managers, registered nurses, enrolled nurses and care workers. One hundred and four staff participated. | - The major themes and sub-themes that emerged from the analysis of the qualitative data included: resident and family; staff; and organization. - Dementia care was related to a lack of recognition of each resident’s care needs and exclusion of family members’ perspectives. Assessment and care planning meant a generalized approach for the residents. - The sub-theme of ‘resident and family centeredness’ related to the recognition of the residents’ needs, inclusion of the voice of residents’ family members in care services; and facilitation of social interactions between residents and their family members. - Most of the care staff identified recognition of needs as an essential part of their daily focus in providing care to the residents. - The quantity and quality of interaction between staff and family members regarding the service and preference of these residents was described by the majority of participants as inadequate. It was conveyed that the perspectives of family members were underrepresented in residential dementia care because it was not a priority for management and staff. Most of the staff described positive outcomes as a result of family members’ visits at aged care homes relating to the mobility, interactions and food intake among residents. - Assessment and care planning emerged as two fundamental aspects of personalised dementia care practice in the views of most staff. - Another aspect to include is the individualised care plan for residents. There were the residents’ needs and wishes to integrate them into care plan. - Most staff considered the positive outcome of non-pharmacological personalised care interventions and found the positive behavioural change of resident. - The second main theme ‘staff’ described a lack of access for staff to ongoing education and training opportunities. - Some staff mentioned their commitment to the routine tasks and how time restriction impacted on their care support and interactions with the residents. | - The experiences and perspectives of the staff provided dimensions of personalised dementia care under the following themes: a requirement of more focus on residents and family members; integrating individualised approach in care assessment and plan; a lack of staff education and training; a lack of person-centered interaction between staff and residents; negative influence of personal and social life of staff in providing care; authoritative leadership and relationships among staff; and a lack of resources and safety. - There was a lack of inclusion of the voice of family members of residents in providing care that impacted on designing the care assessment and plan. The generalised care plan was still in place because of a lack of knowledge and skills among the staff in understanding and managing challenging behaviours for every resident. - A lack of on-going educational opportunity on dementia care in combination with diversity in staff, inability to cope with personalised dementia care and influence of personal life and social issues, all resulted in a sense that personalised dementia care was difficult to maintain in nursing homes. - This study findings indicate that this personalised care practice is yet to be realized because of the challenges associated with shifting from generalized to personalised care. The challenges in shifting to personalised care practice were related to myths/assumptions on behaviours of residents and the associated care burden including workload in the transition of personalised care. - The inclusion of the voice of residents and their family members in designing care plan is in need to establish personalised care practice for residents to give them opportunity to live with a control over care service. - Emphasis on the staff retention and recruitment and resource adequacy is important to improve quality of personalised dementia care in nursing homes. |
| Haugland and Giske (2021) | Norway | Qualitative study | To explore ways of developing appropriate person-centred activities for nursing home residents based on what would be meaningful for them. | Twelve students each year over a three-year period participated in the study (n=36). | - The main theme of this study was ‘Enlivening the residents by cultivating their spark of life’. There were two main categories showing how the nursing home residents became enlivened by sensing the spark of life. - The first category was ‘journeying to meaningful and enlivening (enjoyable) activities’, describing how the students and residents travelled together to search for and adjust to what was meaningful for each resident as the students interpreted the residents’ verbal and nonverbal expressions. - The second category, ‘expressions of enlivening’, showed how students recognized and interpreted the expressions of enlivening among the residents. - The residents with impaired or absent language function expressed themselves using particularly the mouth and eyes. | - The quality of care in nursing homes is measured by several parameters, such as autonomy, social relationships, joy of life, and meaningful activities. - The relational qualities of the nurse-patient interaction are key to the residents’ sense of self-worth and well-being. - Nursing home residents are, to a certain extent, satisfied with the physical care but lack meaningful days, social contact, and the ability to make their own choices. When considering the quality of care in nursing homes, it is crucial to assess all aspects of life that affect health such as the physical, social, mental, and spiritual aspects to understand the person as a complete human being. - To assist the individual in carrying out meaningful activities, the nurse needs to know the biography of the person. - Meaningful activities can simply be an awareness of including the patients in daily activities. - Care must also facilitate meaningful days with the opportunities to make personal choices, engage in social contact, and participate in meaningful activities. - By observing and interpreting what provided the spark of life and enlivened the individuals, they were doing more of the same and observing if the activity still resulted in the same enjoyment. Furthermore, the observations were documented both in the nurses’ logs and in the residents’ journals. By assessing the residents’ past and present interests, nurses consider, safeguard, and strengthen the residents’ resources. - Meaningful activities that gave them the spark of live, the residents remembered the episode much longer than they usually did. - By “journeying with” with the individual resident, the nurse contributes to the resident feeling enlivened, waking up, and experiencing the activity as meaningful. This requires using their creativity, sensitivity, and the try-and-fail approach. |
| Haunch et al. (2023) | UK | Qualitative study | To report on nursing home staff’s perspectives on how and when they meaningfully engaged with residents in the  context of everyday practice. | 21 staff from seven nursing homes. | - Six themes were identified. Four related to how staff engaged with residents with advanced dementia (initiating meaningful engagement, recognising subtle reactions of residents, practising caring behaviour, patience, and perseverance) and two related to when meaningful engagement occurred (lacking time to connect and making the most of time during personal care). - Initiating meaningful engagement: Residents with advanced dementia rarely initiated interactions. Meaningful engagement therefore relied on staff being sensitive to non-verbal signs from residents, such as emotional expression, touch, and posture to know when and how to attempt meaningful connections with residents. Speaking clearly using short sentences, making eye contact, and giving the resident time to respond were techniques used by staff to meaningfully engage. Some care workers described using more physical contact than they did with other residents, such as a pat on the arm and or holding the person’s hand. - Recognising the subtle reactions of residents: An important facilitator of meaningful engagement appeared to be learning to recognise the subtle reactions or behaviours of residents. - Practicing caring behaviours: Staff spoke about the importance of paying attention to their own behaviours in their interactions with residents. Care workers found that approaching residents in a kind, gentle and caring way at a pace that suited the resident resulted in successful engagement with residents. Other caring behaviours included: greeting a person by name or using nicknames, using humour, or sharing information about their own hobbies or family. Patience and perseverance were therefore key to engaging in those special moments of connection. - Lack of time to connect: having the time to connect with residents with advanced dementia was a key concern for staff in this study. While many staff wanted to form connections, they felt they were constantly constrained by having other tasks to do and having other residents to attend to. - ‘Making the most of time’ during personal care: some staff suggested they resolved their main concern of “struggling to find time” by “making the most of time” during personal care. For them, personal care was considered an optimal time to connect. | - Person-centred care (as opposed to person-centred dementia care) is a term used to describe a standard of care that places the person at the heart of care delivery. It can be argued this type of model is not concerned with connections and in contrast, person-centred dementia care highlights the importance of connections and relationship for well-being and the significant role care staff play in ensuring such connections. A significant body research demonstrates that residents with advanced dementia are not commonly in receipt of person-centred dementia care and tend to spend their days alone with little interaction with anyone. - Staff were clear that there was not a single approach to connecting with residents. Those who could adapt, try new things and come back later were most able to describe connections. This relied on initiating meaningful engagement, recognising subtle reactions, practising caring behaviours, being patient and making the most of time during personal care. - The findings suggest personal care as an optimal time to connect. Care workers described their real-world struggles of not having time to connect with people with advanced dementia because of the demands of their work. In this way, they did not need extra time but found a way of capitalising on times of personal care, when they were inevitably spending time close to a resident. - Formalised training consonant with the best aspects of nursing home staff’s existing practice is a possible vehicle to validate good practice and spread it more widely. - Small changes in care workers’ everyday routines can be implemented immediately at little cost and disruption. Emphasising to staff the importance of getting to know residents, learning their communication patterns and using every opportunity as a chance to connect, and making it clear that this is recognised as a core part of their role, could improve meaningful engagement. |
| Ho et al. (2021) | Malaysia | Cross-sectional study | To describe the model of person-centred care adopted by a nursing home, with a logic model and evaluate outcomes on residents’ well-being, care quality, and staff attrition. | Male residents in a 30-bed assisted living facility for persons with dementia. | - Well-Being and Activity Participation: There was a statistically significant improvement in aggregated resident WIB scores in 2016 compared to 2015. Residents were less withdrawn and showed less passive engagement in 2016. - Provision of “comfort” doubled in 2016 compared to 2015, where the incidences of “attachment” in 2016 were 2.25 times more. There was an overall increase in PEs, which contributed to higher resident well-being in 2016. There was a 3.3 times reduction in practices that undermined “inclusion” such as stigmatizing or ignoring. Thus, overall PDs were reduced in 2016 versus 2015. - Staff Turnover Rates: Twenty direct care staff resigned in 2015/2016 as compared to nine in 2016/2017, amounting to 55% reduction in staff turnover rates. | - Patient centred care (PCC) prioritises individual well-being through meaningful occupation and improving the quality of relationships between care provider and recipient. - Today, PCC is synonymous with high-quality care and advocated by many healthcare organisations including the WHO. - Post PCC implementation, the outcomes indicate a superior quality of care, enhanced resident well-being, and better staff retention. - Embracing PCC at every level of the organisation empowered staff to re-invent care with the new priorities of enhancing residents’ well-being, autonomy, and independence. Both classroom and on-the-job training afforded opportunities for role-modelling as an essential component of in-house learning. Having practitioner-leaders steeped in PCC function as mentors and motivators allowed staff to continuously innovate and adapt to the changing needs of the residents, and share strategies that worked. |
| Ibrahim et al. (2020) | Australia | Mixed-methods study | To report on the development and testing of a conceptual model for nursing home care that considers individual residents’ overall needs and the necessary conditions to enable residents to thrive.DS | Stakeholders (n=382) including representatives from nursing homes (nursing staff, residents, family members, executive, board members, therapists and volunteers); industry experts groups and departments; expert informants and committees. | - The model developed consists of five domains relevant to the experience of older adults living in residential aged care— health care; social inclusion; rights; personal care and reablement; and dementia management. - Health care is an obvious inclusion, as many residents have complex health comorbidities and move into nursing homes because of deteriorating health. Health covers both physical health and psychological health. Social inclusion is important, as it is a major contributing factor to quality of life. - Dementia management appears as a domain in its own right given that a large proportion of older adults living in Australian nursing homes have a diagnosis of dementia. | - A nursing home is often the older person's home for several years and therefore must go beyond providing basic health and clinical care to also address individuals’ emotional, social and practical needs. This view is compatible with the tenets of person-centred care, considered the gold standard for care of older people including those with dementia, and with ‘partnering with patients’ in their own care, an important pillar of person-centred care. - Stakeholders agreed that dementia should appear as a domain in its own right once the rationale behind this was explained - a large proportion of residents have a diagnosis of dementia which brings with it unique health-care challenges and makes it a priority area of care. |
| Kemp et al. (2021) | USA | Qualitative study | To outline the range of opportunities for and experiences with meaningful engagement among residents with dementia, and identify successful engagement partner approaches. | 33 residents diagnosed with dementia and their care partners which included nursing home staff (n = 48), external workers (n= 12), volunteers (n= 4), and at least one family member/friend per resident participant (n = 36). | - Residents’ opportunities for and experiences with engagement varied within and across AL communities and among residents over time. Three of the largest communities had formal engagement programing, providing regular opportunities for physical, social, intellectual, and religious activities, including special events and off-site outings. Engagement partners included AL administrative, engagement, care, dietary, housekeeping, maintenance, and transportation staff, volunteers, and co-residents, who were especially significant partners during mealtimes and group activities. We observed meaningful visits from external non-health care workers, including hairdressers, manicurists, and music and pet therapists, and from family members and friends. - Facilitating positive engagement experiences among residents with dementia required observation, attentiveness, and person- and relationship-centered approaches. These approaches included (a) knowing the person (this process involved asking questions, taking an interest in residents’ lives, and collaborating to empower residents to do things for themselves or to do activities that were apt to elicit positive responses. It also meant being observant of individual behaviors and responses), (b) connecting with the person and meeting them where they are (Actively listening and observing verbal and non-verbal cues were keys to connecting and meeting residents on their own terms), (c) being in the moment (An extension of meeting residents where they are, being in the moment demanded attentiveness and adaptability to ongoing change), and (d) viewing all encounters as opportunity (Residents had more frequent and positive outcomes when all interaction partners viewed each encounter as a potential for meaningful engagement. Even small encounters held potential. Care interactions, although routine, represented especially important and oft-overlooked meaningful engagement opportunities. Encounters with non-activity personnel, including kitchen, housekeeping, other staff, and visitors also presented opportunities). | - Meaningful engagement is a critical dimension of quality of life and quality of care for persons living with dementia. - This research confirms the importance of engagement and identifies successful approaches to engaging persons living with dementia in meaningful ways. - Meaningful engagement holds promise for offsetting the negative effects of social distancing for residents and for reducing care partner strain. The successful approaches we identified include (a) knowing the person, (b) connecting with the person and meeting them where they are, (c) being in the moment, and (d) viewing all encounters as opportunity. - These approaches extend other recommendations, including calls to place persons and relationships at the center of care. - We recommend that staff and other care partners receive hands-on, competency-based training on these simple, yet successful approaches with emphasis on maintaining and enhancing relationships with all care partners to support improved quality of life for people living with dementia. Thoughtfully incorporating these approaches into daily life and care routines and adoption by all care partners in resident encounters are ways to maintain and promote meaningful engagement. Whether individual, one-on-one, or group-based engagement experiences, person-centered and relationship-based approaches are foundational to promoting meaningful engagement and improving the quality of life for persons living with dementia in both ordinary and extraordinary times. |
| Laukvik et al. (2022) | Norway | Cross-sectional study | To describe the content and comprehensiveness of nursing documentation for residents living with dementia in nursing homes. | 121 residents in a nursing home with dementia. | - Person-centered content: The life story of the residents was identified in only 15.7% of the reviewed records. Assessment charts, containing information relevant for the patient-centred care (PCC) categories, were identified in 82.6% of the records. 35% of the NDs contained content related to identity and 7% contained information related to inclusion. The identified NDs were most often brief statements about the resident’s general condition. The content of the NDs were most commonly related to pain, behaviour, activity, and family matters. - Comprehensiveness: A Comprehensiveness In Nursing Documentation (CIND) score of 4 was achieved with 31% identified Nursing Diagnosis (NDs) across all PCC categories, meaning that the resident’s problem and corresponding planned interventions were stated, and an effect of the implemented interventions were stated. Only 1% of identified ND achieved a CIND score of 5, meaning that all aspects of the nursing process were recorded, containing descriptions of the resident’s experience. | - Person-centered care (PCC) is considered as high-quality care in dementia long-term facilities, wherein individualised care planning, informed by the residents’ history, needs, and preferences, are recommended. - Unique information about the defining moments in the residents’ lives, and their experiences and beliefs should be provided as a whole in the nursing documentation in order for nurses to relate and interact with the resident in a way that is meaningful and safe. If not reflected in the nursing documentation, it could hinder the healthcare team in accommodating the residents’ individual daily routines, learn about who the residents are and their basic needs. - Structured documentation that demonstrates how the condition of the residents has been understood can contribute to ensuring that they are valued and respected as persons. By connecting information about signs and symptoms, nurses can identify and implement appropriate interventions to achieve desired outcomes. - Increased focus on the perspectives and experiences of the resident in care planning, and documentation of nursing for residents, can create an environment that respects and maintains the selfhood of the resident. - Failure to comprehensively record information related to psychosocial aspects can make it impossible to understand whether important basic needs have been considered in the evaluation processes or whether interventions are appropriate and should be continued. |
| Lee and Bartlett (2020) | UK | Qualitative study | To examine access to, and use of functional objects by people with dementia living in nursing homes, and coin the phrase ‘material citizenship’ to advance thinking and practice in this area. | Thirty-nine participants took part in the study: 15 residents with dementia (11 females and four males, average age 88) 16 staff (12 females and 4 males, average age 46) and eight relatives (six females and 2 males, average age 60). | - Residents who moved directly from hospital to the care home lacked the opportunity to return home to collect their belongings. - Residents were often not involved in deciding which belongings they wanted to take and lacked control over what happened to their items. - None of the participants had an opportunity to return home and had very little control or choice over personal possessions. This indicated a lack of material citizenship. - Staff and relatives reported short timeframes as a barrier to collecting personal possessions. - The responsibility for making decisions and the access people had to functional objects fluctuated between relatives and staff. - There was no evidence of risk assessments taking place concerning functional objects. Instead, staff made subjective decisions on an ad hoc basis. - The local authority care home quality standards, included as a documentary source, provided guidance encouraging certain objects into the care home. The document listed items such as toiletries, soft furnishings, photographs and a small item of furniture. Clothing should be able to withstand a 60-degree wash, be a size bigger than usual and have an elasticated waistband to assist staff with dressing and undressing. Objects were viewed and controlled through a service lens rather than as items important for enabling citizenship, or as a mechanism for gaining a deeper understanding of a person's historical biography and present identities. | - People with dementia are routinely excluded from the decision-making process. In the context of this study, functional objects manifest in risk-averse practices. While risk management is necessary in care provision and can be a positive process, it can also disempower and be restrictive if done so discriminatorily. Examining decision-making and the use of objects is useful in drawing out care practices that do or do not afford people with dementia the same opportunities as others. - Residents’ interactions with their material surroundings provide the ability to continue routine practices such as organising their surroundings, cleaning and hosting. Staff however were often observed discouraging interactions from taking place. These were subtle injustices in care practice. Injustices that were not intentional, but nevertheless present. - Functional objects are overlooked in the context of care homes and there is a lack of guidance for care workers. - Understanding how functional objects manifest in care practices made visible the lack of social citizenship afforded to people with dementia in this setting. It would be valuable for care practices to combine a material citizenship approach with existing care practices. This would elevate the importance of object–person relations, allowing staff and relatives to recognise that functional objects can help a person maintain identity and influence how they are perceived. |
| Malta-Müller et al. (2020) | Denmark | Qualitative study | To explore how everyday life was organised in a Danish nursing home for people living with advanced dementia and how relatives experienced their family members’ everyday lives. | Field notes of observations of a nursing home. Participants included 9 relatives of nursing home residents. | - Enabling a meaningful everyday life was identified as a main theme with two sub-themes: (1) Structures of daily life: Balancing collective and individual activities and (2) Physical togetherness: Balancing being together and being alone. - Everyday life in the nursing home followed the same overall structure organised and maintained by leaders and other staff. Overall, relatives experienced that their family members were thriving in the nursing home. This was particularly related to staff members’ facilitation of daily activities. Collective activities ensured that something happened in the residents’ everyday lives and this was perceived as an important feature of the care. Such experiences served as an argument in favour of the more structured and active everyday life experienced in this nursing home. Despite the daily structure, there was some level of flexibility within the regularity. For instance, not all residents participated in the collective activities on a daily basis. Thus, although staff members encouraged participation, they would also accept residents’ declining to participate. In this way, staff tried to achieve a balance between maintaining structure and safeguarding individual needs and wishes. - Balancing being together and being alone: The structures of daily life entailed that physical togetherness was an unavoidable part of everyday life in the nursing home, and especially the presence of staff was an important reason for residents to reside in the shared living room. Thus, relatives viewed the physical closeness of staff as a way of ensuring the ability to respond to each resident’s needs whenever required. In this way, background knowledge about the individual resident was used to support personal well-being. | - In Denmark, a research project recently explored the care in a dementia-specific municipal day care unit and nursing home known for its good reputation. The findings showed that enabling a good everyday life for users and residents entailed staff creativity and initiation of activities that seemed meaningful in the moment. - The findings from this study showed that activities and togetherness were used by leaders and other staff in an effort to enable a meaningful everyday life for residents living with advanced dementia in the selected nursing home. Overall, relatives believed that this organization contributed to a good life for their family members. However, both features entailed a continuum, and staff became important facilitators of a suitable balance between the outer poles. This balance could sometimes be challenging to maintain for all residents at the same time. |
| Martinsson et al. (2020) | Sweden | Cross-sectional study | To investigate whether quality of end-of-life care for patients with dementia depends on age, gender and place of death. | A total of 4,752 with Alzheimer’s disease and 11,710 with other or unspecified dementia, were included in the study. | - When not adjusting for age or gender, 10 of the 13 measured end-of-life care outcomes showed better outcomes in nursing homes compared to hospitals, while 2 outcomes were better in the hospital group. - Giving information to next of kin about the transition to end-of-life care was more common in hospitals, but offering a follow-up talk to next of kin after death of the patient was more common in nursing homes. - Patients who died in nursing homes were more likely to be assessed for pain and other symptoms during the last week of life, to be prescribed drugs against pain, nausea and anxiety, and to have someone present at the moment of death compared to patients who died in hospitals. - Pressure ulcers at the time of death were more common in hospitals. | - A majority of patients with dementia were given end-of-life care in the nursing home, indicating that a majority of patients suffering from dementia can be managed in nursing homes during end of life. - Death in hospitals was associated with poorer quality of end-of-life care compared to death in nursing homes. - Study data support the importance of advance care planning and individual assessments in nursing homes to avoid referral to hospitals during end of life. Despite established recommendations to avoid hospitalisation if possible, there were strong associations between younger age, male gender and hospitalisation in the end of life. |
| Nygaard et al. (2021) | Norway | Qualitative study | To examine what matters to nursing home residents with dementia by exploring their perceptions of nursing home health care through the conceptual lens of person-centred care. | 35 nursing home residents all diagnosed with dementia participated. | One overarching theme and four interrelated subthemes emerged.  The overarching theme was:   - Different matchings of person-centred care and routines in health care   The subthemes were:   - Understanding of the interplay between disabilities and ageing: the residents missed the feeling of being an individual with their own preferences and needs and missed personalised and tailored PCC. They talked about their previous lives as being part of their identity, indicating that it was important for health-care providers to have knowledge of their past to understand and support their current needs. They reported, however, that the health-care providers seemed to forget the person behind the dementia disease and their age, expressing that their feelings and experiences of being affected by dementia and living in a nursing home in general were not taken into account. - Participating based on one's own preferences and needs: the residents appreciated engaging in their own care. Some highlighted the importance of opportunities to participate in daily tasks based on their needs and abilities. It was important for the residents to decide when to get up in the morning, when to go to bed, whether to enjoy alcohol or which clothes to wear, for example. Their statements indicated that they were required to follow the nursing home's routine and that their own preferences were seldom taken into account. The residents felt that it was important to have some control and choice regarding everyday life. - Incongruence between the person with dementia's preferences and needs and health-care support: It seemed important that the health-care providers combine interpersonal competences and practical skills in their daily practice to meet the complex and very different preferences and needs of the residents. Incongruence between the residents’ preferences and the health care they received had an impact on well-being. - Working conditions: the relationship between residents and healthcare providers: Several residents highlighted the importance of developing a relationship with and confidence in the health-care providers, as well as health-care providers’ knowledge about each resident. The residents observed that the health-care providers were busy, so they did not want to disturb them unnecessarily. | - Health care in nursing homes must follow a more person-centred approach, even though some residents expressed satisfaction with the health care they received. - Even though there is a consensus about the relevance of a person-centred perspective, a translation of the PCC framework into practice is needed, particularly in nursing homes, where the coexistence of disability, cognitive decline, and chronic conditions, challenge resident’s everyday lives and the provision of care. - Organisational conditions can reinforce a task-oriented care approach, which is generally viewed as the opposite of a patient oriented approach. Finding a balance between organisational and administrative routines and the expectations of older people's needs can facilitate individualised care. - The main challenge regarding health-care quality in nursing homes is finding a balance between the general routine and the residents’ individual preferences and needs. Despite health-care providers’ substantive focus on PCC, the results show that nursing home residents are asking for more PCC. Health care in nursing homes should enable residents to participate in daily activities, support decision-making and sustain their personhood. - The practices of health care should consider including specific professional approaches to build relationships and engage residents in activities. The results indicated that health care in nursing homes must follow a more person-centred approach allowing the negotiation of health care based on what matters to the residents. Healthcare providers should give residents a voice while caring. |
| Peoples et al. (2018) | Denmark | Qualitative study | To explore the way in which relatives of people with dementia and healthcare professionals create and maintain a meaningful everyday life for the residents in a Danish dementia village. | 32 participants took part in the study. The relatives consisted of three daughters, one son, two siblings, two spouses, one of whom was a widower to a former resident. The staff consisted of certified healthcare assistants and helpers. | - One main theme, ‘Enabling a familiar and meaningful everyday life in the dementia village’ with three corresponding subthemes: (1) The everyday life is what matters, (2) The importance of still being part of the ‘real’ life outside and (3) Using the opportunities in the dementia village, were identified. - The everyday life is what matters: Relatives and staff described how they individually and collaboratively worked to enable a meaningful everyday life for the residents. For the relatives it was important to engage in familiar activities they used to share with the residents and that used to characterise their relationship based on their role as an adult child, a sibling or a spouse. The staff also emphasised the importance of enabling a meaningful everyday life for the residents, but could at times have a different perspective of what this entailed, for instance in regard to the daily activities. - The importance of still being part of the ‘real’ life outside: several relatives felt that it was important for the residents to maintain a connection to the life outside the dementia village and they therefore prioritised to enable this. - Using the opportunities in the dementia village: Many of the relatives and staff had witnessed the transformation from a regular nursing home to a dementia village and all the changes this had resulted in, for instance the creation of a small ‘street’ with a shop, a kitchen, a hairdresser, a fitness room, a music library and a ‘man cave’, and they primarily experienced these changes as positive. | - The findings indicate that a more systematic and collaborative way for relatives and staff of working together to create meaningful and joyful moments in the day-to-day life is needed, for instance through systematic use of life stories. The relatives are able to portray the person they know and love, and provide information about former meaningful activities of the residents and the staff are able to provide professional knowledge about ways to adapt these activities to the resident’s current condition as the illness progresses. - The findings in this study support the need for creating environments that provide various possibilities of engagement in meaningful activities in the outdoor as well as indoor environment in order to enhance the overall quality of the lives of all the residents. However, the findings also indicate that in order for residents with advanced dementia to take advantage and benefit from the possibilities provided in the dementia village, more staff and volunteers are needed. - The findings indicated that both groups were committed to create a meaningful everyday life for the residents, but also revealed different meanings of when, where and how a meaningful everyday life could be understood and best be achieved. The findings showed that people with advanced dementia were not able to take advantage and benefit from the activities and possibilities that were provided in the dementia village, since this required support beyond what could be provided. |
| Pruitt (2022) | USA | Cross-sectional study | To determine the impact of screening for changes and physical function on the number of palliative care referrals among people with dementia living in nursing homes. | A purposive sample of nursing home residents who have been diagnosed with dementia was used for this project. | - Gender plays a significant role in determining whether an order for palliative care is received. Specifically, female residents are more likely to get a palliative care order relative to their male counterparts. - The facility was noted to have several floors including a specialized secure dementia unit. Although, residents were specifically noted as having dementia in the special care unit, it was not statistically significant, p = 0.170. - Analyses conducted for falls with serious injury yielded a statistically significant relationship (p = 0.041) with obtaining orders for palliative care. In like manner, visits to the emergency department yielded comparable results, p = 0.02, and indicated a very significant relationship with obtaining orders for palliative care. Decreased oral intake, p =0.034, was statistically significant in obtaining a palliative care order. If a resident with dementia has decreased oral intake, the likelihood of that resident receiving an order for palliative care is increased. The most statistically significant relationship was noted between decreased physical activity and obtaining an order for palliative care, p < 0.001. | - The palpable loss of administrative staff and use of interim and temporary staff for administrative and clinical positions undermined the consistency needed in providing care, thereby reducing the quality of care being rendered. - Residents living with dementia in long-term care facilities are challenged with change daily. The idea that each day is new, and a person may not remember all, or parts of their previous life can lead to challenges that may not be identifiable to short term staff. Over extended staff may also miss key indicators due to apathy, fatigue, or feeling overworked that can adversely affect persons living with dementia in nursing homes. The ability of staff to identify deconditioning and changes that occur with this population is a key component in helping residents to live their life to the fullest potential. - The challenges with staffing also impacted the staff education component, including time, the availability of educators and staff to provide and receive the education. The results of this project align with the literature in the use of a reliable palliative care screening tool for patients living with dementia can positively affect patients. |
| Rivera-Hernandez et al. (2022) | USA | Cross-sectional study | To determine whether disparities among Medicare beneficiaries living with dementia exists in five different long-stay quality measures. | A total of 1,005,781 long-term residents from 15,137 nursing homes. | - All minority groups were 6% less likely to go to nursing homes with an Alzheimer’s unit and facilities which are for profit compared to White residents. Similarly, White residents went to facilities with a high proportion of residents who are from the same racial group (85%). - Compared to White, Asian/Pacific Islander and Hispanic residents were slightly more likely to live in nursing homes with a higher proportion of people with severe cognitive impairment. American Indian/Alaska Native residents were more likely to reside in facilities with a higher proportion of residents that reported daily pain (10%) compared to White (7%) residents. - Black and Hispanic residents were living in nursing homes with 2% higher readmission rates and they were at least 8% less likely than White residents to go to nursing homes with high-quality ratings. - White residents reported higher use of antipsychotic medications along with Hispanic and American Indian/Alaska Native residents (28%, 28%, and 27%, respectively). Yet Asian/ Pacific Islander and Hispanic residents reported slightly higher percentages of physical restraint usage compared to White residents. | - There were major disparities in access to quality care facilities by minority groups. - As CMS continues to increase quality among long-term care residents post-COVID-19 pandemic, a major emphasis needs to be placed on eliminating disparities in access to high-quality nursing homes and better long-term care options among minority elders. |
| Rosenthal et al. (2020) | USA | Qualitative study | To investigate health professionals’ experiences with decision-making during changes under the National Partnership to  Improve Dementia Care in Nursing Homes. | 40 nursing home staff (primarily nursing, activities, and social services staff), and 10 prescribing physicians. | - Five recurring themes relating to decisions during changes in antipsychotic medication use since 2012 were identified: (a) staff and physicians are aware of the need to reduce antipsychotic medication use; (b) the value of person-centered approaches to accomplish these reductions; (c) the contribution of collaboration and communication to achieving reductions; (d) the need for more training and education about dementia and for more staffing; (e) the challenges posed by CMS regulations and surveys. - Respondents were very aware of the need to reduce antipsychotics, for the well-being and safety of residents. Respondents were aware of state and national rates of antipsychotic medication use, in comparison to the state average because it impacted their standing in relation to nearby nursing homes. This awareness was due in part to external pressures, particularly from state nursing home surveyors, who pay close attention to antipsychotic medication rates. Pressure also stemmed from within facilities. However, some staff members suggested that medications address aggressive behaviours of residents more quickly. - Individualised, person-centered approaches were deemed essential to reducing use of antipsychotics. To accomplish this, prescribers and non-prescribers described several steps, including ruling out medical causes of difficult behaviours, and trying nonpharmacological alternatives first, as well as non-antipsychotic drugs, such as mood stabilizers. - Respondents discussed the need for additional resources: nonpharmacological alternatives require more time and an individualised approach compared to the use of medications, which eliminate symptoms rapidly. - Survey’s Respondents acknowledged CMS regulations helped spur quality improvement efforts. But they often felt the survey process was “too black and white,” with surveyors often too focused on “the number,” while overlooking complexities of reducing antipsychotics. | - Respondents emphasised the value of person-centered approaches, consistent with research on a variety of nonpharmacological interventions. - Responses confirmed the concerns in other studies that greater knowledge, education, and training are needed for nursing home staff, prescribers, and residents’ families about causes and behaviours in dementia, and its safe and effective management. - Respondents indicated that patient-centered care cannot be done by formula, but they expressed confidence that the medication reduction can be done on a case-by-case basis. These responses point to the need for additional resources for training and staffing, if nursing homes are to provide the optimal level of services to address dementia without antipsychotics. - As a result, the practice implications include continuing training of all staff, team review of cases with antipsychotic prescriptions, and establishing a focus with families, prescribers, and the whole team on safer dementia care. Nursing home administrators who want to succeed in this effort will note that training can be time-consuming, but online training programs in dementia for direct care and non-direct care workers can help reduce staff burdens and potentially improve the lives of residents with dementia. |
| Royston et al. (2020) | UK | Mixed-method study | To describe the methodology behind the dementia care framework and outcomes data from the first phase (of 20 care homes that included the care of 451 people living with dementia). | 20 care homes throughout the UK. 13 from England, 4 from Scotland and 3 from Northern Ireland. 451 residents living with dementia lived across these 20 care homes. In total, 538 colleagues directly cared for people with dementia care. | - The dementia care framework helped care teams identified 8,176 specific individual elements of care based on the individual care reviews. - The most common areas that required improvement were that the care plan identifies: - any spiritual preferences and practices that the resident may have and details of how these needs are met - details of what items/people the resident has formed attachments to and how they feel when separated from them - details of what is psychologically comforting for the resident (e.g. appropriate use of touch etc.) - details of how the resident's occupational needs (e.g. vocational activities) are met - those community links that are important to the resident and shows how staff facilitate these - details of how to maintain and support the resident's unique identity - how the resident wishes to be included in communal activities - evidence that their choices are respected - if there is a lasting power of attorney or enduring power of attorney a copy of this is held | - As noted in the results section, many of the aspects of which improved related to a resident’s psychological, social and spiritual needs. Good physical care and support of a person’s medical needs can overlook other broader holistic needs. These broader holistic needs are important to consider as they contribute to enhancing a person’s overall wellbeing. - Implementation of the dementia care framework helped enhance the personhood of people living with dementia within care homes as it sought to illuminate aspects of life which may had diminished like; engagement with one’s local community, or fulfilment of one’s spiritual need or indeed provision and engagement in meaningful modes of life and activity. |
| Steele et al. (2020) | Australia | Qualitative study | To examine understanding of confinement by exploring the daily facilitators of confinement in the lives of people with dementia. | Five people living with dementia in care homes, 19 care partners, 12 care home professionals, and 9 lawyers and advocates. | - Many participants identified material features of restraint that restrict the movement of people living with dementia. However, participants generally mentioned these features only after being specifically asked about them, suggesting that they are unquestioned aspects of care home culture. - Some care partner participants had knowledge of chemical restraint in relation to their spouses or parents. - Restrictive practices are used in care homes and affect mental well-being. - The most commonly mentioned factor affecting freedom of movement was the removal of means of mobility. This included not providing mobility aids, opportunities for physical exercise, or meaningful activities to prevent decline and distress. - While some care partners spoke positively of the recreational and social opportunities available to residents, some participants noted that people living with dementia were not always included in the full range of external activities on offer. - Care home professionals and care partners noted that the lack of understanding of dementia in the community caused fear, which could lead to residents being prevented from accessing the community. - Participants noted that public spaces outside of the care home might not be physically accessible for people living with dementia. | - Cultural logics around dementia that sustain confinement include the logic is that dementia is feared and needs to be hidden from the community. - Another logic is that people living with dementia lack their own subjectivity and ability to articulate their needs and experiences. All of these logics speak to a profound tolerance of the inequality and dehumanisation of people living with dementia. - While participants mentioned physical and environment restrictions on liberty (predominantly locked doors and gates) and ways in which people living with dementia had limited or no access to community activities or spaces, they generally did not perceive these as forms of confinement or restrictions on liberty or, indeed, even as problematic or unusual. This suggested that those involved in the day-to-day support and advocacy of people living with dementia are largely oblivious to significant human rights violations. - People living with dementia are effectively fenced in by both the stigma around dementia and the staff perception that negative community experiences are too difficult to manage. Yet, at the same time, the segregation of people with dementia cuts off opportunities for the whole community to develop a better understanding of living with dementia, thus reinforcing the “need” for confinement. |
| Sund et al. (2023) | Norway | Qualitative study, synthesis of ethnographic stories. | To show how older residents living with dementia in nursing homes can realise their everyday citizenship. | Two nursing homes in Norway. | - Staff expressed deep caring about their wellbeing. Laughter, sit-down conversations and a quest for both calm and engagement in activities was observed. Staff stated their aspiration for person-centred care. - An atmosphere of waiting sometimes emerged, such as waiting for meals. Even though activities and events differed from day to day, nursing home life relied on a set structure and regular themes. Meals emerged as a predictable frame and acted as a reference point for staff and residents. Residents gathered at mealtimes and had pre-determined seats, but often sat in silence. Several residents expressed joy or anticipation when invited to events and activities or receiving visits from children and others in the local community. Other times residents closed their eyes or slept on and off during the day in the common areas, mostly when there was nothing going on or no one to whom to talk. | - Within everyday environments of care, spontaneous initiatives and agency, working towards contribution and responsibility within mundane and habitual everyday doing, promotes citizenship. - Becoming adds to the practical understanding of citizenship through its processual perspectives of human occupation, within repetitive and cumulative moments of change, engagement, responsibility or contribution. - Everyday life perspectives within professional practice for persons living in nursing homes is important. A citizenship of becoming presupposes that institutional perceptions of activities as offered ought to be broadened towards supporting residents’ natural desires to do and act within the mundane and ordinary of everyday life. |
